# Large-scale Mendelian randomization identifies novel pathways as therapeutic targets for heart failure with reduced ejection fraction and with preserved ejection fraction

**DOI:** 10.1101/2024.03.22.24304728

**Authors:** Danielle Rasooly, Claudia Giambartolomei, Gina M. Peloso, Hesam Dashti, Brian R. Ferolito, Daniel Golden, Andrea R. V. R. Horimoto, Maik Pietzner, Eric H Farber-Eger, Quinn Stanton Wells, Giorgio Bini, Gabriele Proietti, Gian Gaetano Tartaglia, Nicole M. Kosik, Peter W. F. Wilson, Lawrence S. Phillips, Patricia B. Munroe, Steffen E. Petersen, Kelly Cho, John Michael Gaziano, Andrew R. Leach, VA Million Veteran Program, John Whittaker, Claudia Langenberg, Nay Aung, Yan V. Sun, Alexandre C. Pereira, Jacob Joseph, Juan P. Casas

## Abstract

We used expression quantitative trait loci (eQTLs) and protein quantitative trait loci (pQTLs) to conduct genome-wide Mendelian randomization (MR) using 27,799 cases of heart failure (HF) with reduced ejection fraction (HFrEF), 27,579 cases of HF with preserved ejection fraction (HFpEF), and 367,267 control individuals from the Million Veteran Program (MVP). We identified 70 HFrEF and 10 HFpEF gene-hits, of which 58 are novel. In 14 known loci for unclassified HF, we identified HFrEF as the subtype responsible for the signal. HFrEF hits *ZBTB17*, *MTSS1*, *PDLIM5*, and *MLIP* and novel HFpEF hits *NFATC2IP,* and *PABPC4* showed robustness to MR assumptions, support from orthogonal sources, compelling evidence on mechanism of action needed for therapeutic efficacy, and no evidence of an unacceptable safety profile. We strengthen the value of pathways such as ubiquitin-proteasome system, small ubiquitin-related modifier pathway, inflammation, and mitochondrial metabolism as potential therapeutic targets for HF management. We identified *IL6R*, *ADM,* and *EDNRA* as suggestive hits for HFrEF and *LPA* for HFrEF and HFpEF, which enhances the odds of success for existing cardiovascular investigational drugs targeting. These findings confirm the unique value of human genetic studies in HFrEF and HFpEF for discovery of novel targets and generation of therapeutic target profiles needed to initiate new validation programs in HFrEF and HFpEF preclinical models.

## 1. INTRODUCTION

Heart failure (HF) is classified into two major clinical subtypes based on the assessment of contractility as measured by the left ventricular ejection fraction (LVEF): HF with reduced ejection fraction (HFrEF), defined as LVEF less than or equal to 40%, and HF with preserved ejection fraction (HFpEF), defined as LVEF greater than or equal to 50%^1^. While significant advances in the therapy of HFrEF have improved outcomes over the last three decades, the morbidity and mortality contributed by this systemic condition remain high^2^. Additionally, treatment for HFpEF remains a major unmet global need, and with the exception of sodium glucose cotransporter 2 (SGLT-2) inhibitors that have been proven beneficial in HFpEF and HFrEF^3^, most drugs that have been tested for in HFpEF phase 3 trials have not shown a clear benefit^4,5^. The therapeutic challenge of HFpEF is not surprising, considering the large difficulties in developing preclinical models that can faithfully reproduce human HFpEF, a heterogeneous phenotype^6^.

Human genetics is a well-established strategy for identifying drug targets and causal pathways that can further inform drug discovery. Except for a genome-wide association study (GWAS) in HFrEF and HFpEF from the Million Veteran Program (MVP)^7^ conducted by our group^8^, most loci that have been identified have been derived from genetic studies on unclassified HF^8–10^. Despite its value in uncovering novel mechanisms in HF, the lack of genetic evidence specific to HFrEF and HFpEF presents a challenge in developing and validating preclinical models and in selecting drug targets to test.

To overcome these limitations, we conduct a genome-wide Mendelian Randomization (MR) study that leverages MVP Release 4 GWAS data using 27,799 HFrEF and 27,579 HFpEF^11^ cases with protein and expression quantitative trait loci (pQTLs and eQTLs). Specifically, we use pQTLs from the SOMAscan V4 assay covering 5,207 aptamers capable of measuring 4,988 unique human proteins available in deCODE^12^, Fenland^13^, and Atherosclerosis Risk in Communities (ARIC)^14^ studies, and eQTLs from GTEx V8^15^ and eQTLGen^16^. For every gene identified as a hit by MR, we triangulate MR findings with orthogonal sources of evidence to strengthen causal links from proteins to disease and create a therapeutic target profile covering efficacy, on-target safety, novelty of biological mechanism, druggability, and predicted mechanism of action (MoA) needed for a therapeutic solution. Finally, we conduct replication of hits using multi-ancestry datasets from BioVU and MVP containing 12,604 HFrEF cases, 11,486 HFpEF cases, and 151,118 control individuals of African-American, Hispanic, and European-descent without history of any HF, and an alternative platform replication using the antibody-based Olink assay.

## 2. RESULTS

### General findings

Figure 1 illustrates the overall design. Of 15,253 evaluated protein-encoding genes, we identified ten genes for HFpEF and 70 genes for HFrEF that passed our Bonferroni correction threshold (*p*-value < 2.06 × 10^−6^), including the cardiomyopathy-associated genes *BAG3* and *TCAP*^17^. To define novelty of hits, we compared our findings against all previously published GWAS and MR studies in HF and HF subtypes^8,9,18–23^. For HFpEF, nine genes are novel, and one is a replication of prior reports^8,9,18–23^, while for HFrEF, 49 are novel, 14 have not been previously reported for a specific HF subtype (“partially novel”), and seven are replications of prior reports ^8,9,18–23^; see **Table 1** and **Tables S1** to **S3**. At a false discovery rate (FDR) of 5% (*p*-value < 6.80 × 10^−4^), a total of 49 genes for HFpEF (10 of which also met Bonferroni) and 362 genes for HFrEF (70 of which met Bonferroni) passed our criteria for suggestive hits. **Figure S1** illustrates the overlap between our hits and previously reported hits. As suggestive hits, we identified known cardiomyopathy genes *ACTN2*, *FLNC*, *RIT1*, and *SGCD*. No single gene passed the Bonferroni correction threshold for both subtypes. However, *FTO* was a hit for HFpEF and a suggestive hit for HFrEF, and *MYOZ1 was a* hit for HFrEF and a suggestive hit for HFpEF (**Table S2**). The genes with the strongest odds ratios (OR) against HF subtypes were *ZBTB17*, *ERBB2*, *LAG3*, and *TCAP*, all with OR higher than 1.5, and *SORT1*, *TNFRSF6B*, *TCTA*, *C3orf62*, *PSRC1*, and *LTN1*, all with OR lower than 0.67, suggesting a strong protective effect.

**Figure 1.**
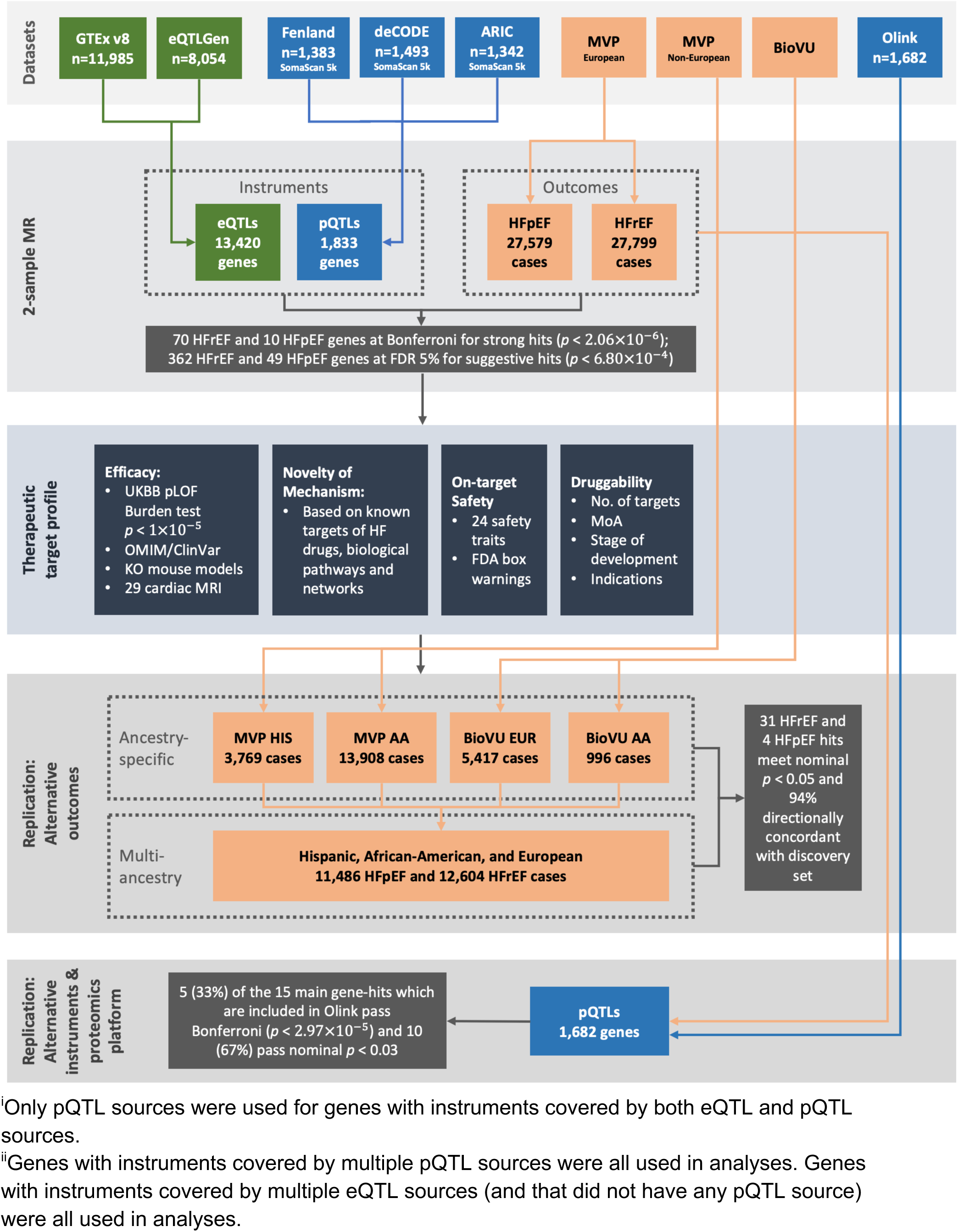
Schematic of study design. We evaluated the putative causal effect of eQTLs from GTEx and eQTLGen and pQTLs from Fenland, deCODE, and ARIC on HFpEF (27,579 cases) and HFrEF (27,799 cases) in a cohort of 422,645 individuals from the Million Veteran Program (MVP) using two-sample Mendelian randomization (MR). We evaluated 11,985 genes from GTEx v8 and 8,054 genes from eQTLGen, totaling 13,420 unique genes among the two eQTL sources. For the pQTL sources, we evaluated 1,393 genes from Fenland, 1,493 genes from deCODE, and 1,342 genes from ARIC, totaling 1,833 unique genes among the three pQTL sources. Primary hits are identified using a Bonferroni-adjusted threshold of *p*-value < 2.06 × 10^−6^ and suggestive hits are identified using a false discovery rate (FDR) of 5% (*p*-value < 6.80 × 10^−4^). For all hits, we created a therapeutic target profile consisting of efficacy, novelty of mechanism, on-target safety, and druggability annotations. We conducted ancestry-specific replication leveraging data from Hispanic [HIS] and African-American [AA] ancestry individuals in the MVP and European [EUR] and African-American [AA] ancestry individuals in the Vanderbilt BioVU, and multi-ancestry replication on 175,208 individuals using a meta-analyzed GWAS for HFpEF (11,486 cases) and HFrEF (12,604 cases). We conducted an instrument and proteomics platform replication using plasma proteomic profiles of 54,219 individuals using the antibody-based Olink Explore 3072 platform data generated by the UK Biobank Pharma Proteomics Project.

**Table 1.** Novel and partially novel Mendelian randomization (MR) hits for HFrEF and HFpEF in the Million Veteran Program (MVP) Release 4. Novel hits are those that have not been previously reported as a hit by published GWAS or MR studies for unclassified HF, HFrEF, or HFpEF^8,9,18–23^. Partially novel hits are those that have been previously reported as a hit for unclassified HF by published GWAS or MR studies. Gene names that are marked with an asterisk have been found significant (*p*-value < 2.06 × 10^−6^) in multiple data sources or tissues in the MR analyses. For each gene, if multiple MR sources (pQTL studies or eQTL tissues) were available, we report on the finding with the lowest MR *p*-value. Findings are ordered by the lowest MR *p*-value.

| Protein Gene Name | HF Subtype | Source | # SNPs | Odds ratio | 95% CI | p value | MR Egger intercept; p value | Q p value |
| --- | --- | --- | --- | --- | --- | --- | --- | --- |
| **Novel hits** |  |  |  |  |  |  |  |  |
| *LAG3* | HFrEF | deCODE | 2 | 1.77 | (1.37,2.17) | 6.18E-07 |  | 0.56 |
| *TCAP* | HFrEF | GTEx Muscle Skeletal | 1 | 1.62 | (1.32,1.92) | 2.80E-07 |  | NA |
| *ERBB2\** | HFrEF | GTEx Skin Not Sun Exposed Suprapubic | 1 | 1.57 | (1.36,1.78) | 5.12E-11 |  | NA |
| *ATP6V1F* | HFrEF | GTEx Skin Not Sun Exposed Suprapubic | 1 | 1.49 | (1.25,1.74) | 1.97E-06 |  | NA |
| *SENP3* | HFrEF | GTEx Esophagus Mucosa | 1 | 1.38 | (1.22,1.55) | 9.92E-08 |  | NA |
| *RNF123* | HFrEF | GTEx Skin Sun Exposed Lower leg | 1 | 1.38 | (1.2,1.55) | 1.24E-06 |  | NA |
| *KLHDC8B\** | HFrEF | GTEx Artery Tibial | 1 | 1.35 | (1.21,1.49) | 3.28E-08 |  | NA |
| *MED1* | HFrEF | eQTLGen | 1 | 1.34 | (1.18,1.5) | 1.36E-06 |  | NA |
| *MST1R* | HFrEF | GTEx Lung | 1 | 1.30 | (1.16,1.43) | 9.64E-07 |  | NA |
| *WDR6\** | HFrEF | GTEx Colon Sigmoid | 1 | 1.28 | (1.17,1.4) | 4.73E-08 |  | NA |
| *QRICH1\** | HFrEF | GTEx Esophagus Gastroesophageal Junc | 1 | 1.27 | (1.16,1.39) | 6.00E-08 |  | NA |
| *SEC24C* | HFrEF | GTEx Esophagus Mucosa | 1 | 1.26 | (1.14,1.37) | 9.27E-07 |  | NA |
| *RHOA* | HFrEF | GTEx Muscle Skeletal | 1 | 1.25 | (1.15,1.36) | 7.98E-08 |  | NA |
| *CA9\** | HFrEF | GTEx Artery Tibial | 1 | 1.25 | (1.14,1.36) | 2.32E-07 |  | NA |
| *INPP5F* | HFrEF | GTEx Cells Cultured fibroblasts | 2 | 1.23 | (1.13,1.33) | 3.13E-07 |  | 0.00 |
| *NFATC2IP* | HFpEF | GTEx Skin Not Sun Exposed Suprapubic | 3 | 1.21 | (1.13,1.29) | 7.45E-09 | 0.03; 0.85 | 0.97 |
| *ZNF664* | HFrEF | GTEx Thyroid | 1 | 1.18 | (1.11,1.26) | 2.01E-07 |  | NA |
| *STARD3* | HFrEF | GTEx Cells Cultured fibroblasts | 1 | 1.17 | (1.11,1.23) | 6.13E-10 |  | NA |
| *GMPPB* | HFrEF | GTEx Testis | 1 | 1.16 | (1.09,1.23) | 1.79E-06 |  | NA |
| *MMP11\** | HFrEF | GTEx Heart Atrial Appendage | 1 | 1.16 | (1.09,1.22) | 7.00E-07 |  | NA |
| *ATF1\** | HFrEF | GTEx Colon Sigmoid | 1 | 1.15 | (1.1,1.21) | 2.65E-08 |  | NA |
| *CYP1A1\** | HFrEF | GTEx Adipose Visceral Omentum | 1 | 1.14 | (1.09,1.19) | 3.67E-08 |  | NA |
| *CCDC71\** | HFrEF | GTEx Brain Cerebellum | 1 | 1.12 | (1.07,1.16) | 5.01E-08 |  | NA |
| *MRPL35* | HFrEF | GTEx Artery Aorta | 2 | 1.09 | (1.05,1.13) | 8.79E-07 |  | 0.87 |
| *MTSS1* | HFrEF | GTEx Heart Left Ventricle | 1 | 1.09 | (1.06,1.12) | 2.21E-09 |  | NA |
| *ARL17A\** | HFpEF | GTEx Brain Amygdala | 1 | 1.06 | (1.04,1.09) | 6.27E-07 |  | NA |
| *LRRC37A2\** | HFpEF | GTEx Cells Cultured fibroblasts | 1 | 1.06 | (1.03,1.08) | 1.39E-06 |  | NA |
| *LRRC37A* | HFpEF | GTEx Skin Sun Exposed Lower leg | 2 | 1.05 | (1.03,1.07) | 2.07E-08 |  | 0.23 |
| *SORT1\** | HFrEF | GTEx Liver | 1 | 0.95 | (0.93,0.97) | 1.04E-08 |  | NA |
| *PROM1\** | HFrEF | GTEx Artery Aorta | 1 | 0.95 | (0.93,0.97) | 1.90E-06 |  | NA |
| *MAPT* | HFpEF | GTEx Colon Sigmoid | 2 | 0.94 | (0.92,0.96) | 3.31E-09 |  | 0.63 |
| *PDLIM5* | HFrEF | GTEx Cells Cultured fibroblasts | 2 | 0.92 | (0.89,0.95) | 1.92E-07 |  | 1.00 |
| *CCDC36\** | HFrEF | GTEx Prostate | 1 | 0.92 | (0.89,0.95) | 1.84E-08 |  | NA |
| *SENP7* | HFrEF | eQTLGen | 1 | 0.92 | (0.88,0.95) | 2.03E-06 |  | NA |
| *NCKIPSD\** | HFrEF | GTEx Brain Cerebellar Hemisphere | 1 | 0.91 | (0.88,0.95) | 2.89E-07 |  | NA |
| *CCDC92\** | HFrEF | GTEx Heart Left Ventricle | 1 | 0.88 | (0.85,0.92) | 3.61E-08 |  | NA |
| *AMT\** | HFrEF | GTEx Artery Tibial | 1 | 0.88 | (0.84,0.92) | 3.46E-08 |  | NA |

| Protein<br>Gene<br>Name | HF<br>Subtype | Source | # SNPs | Odds<br>ratio | 95% CI | p value | MR Egger<br>intercept;<br>p value | Q p value |
| --- | --- | --- | --- | --- | --- | --- | --- | --- |
| **Novel hits (con't)** |  |  |  |  |  |  |  |  |
| *DNAH10\** | HFrEF | GTEx Colon Sigmoid | 1 | 0.88 | (0.84,0.92) | 2.00E-07 |  | NA |
| *MPI* | HFrEF | GTEx Spleen | 1 | 0.88 | (0.84,0.92) | 3.47E-08 |  | NA |
| *ARIH2\** | HFrEF | GTEx Adrenal Gland | 1 | 0.86 | (0.82,0.91) | 3.28E-08 |  | NA |
| *P4HTM\** | HFrEF | GTEx Brain Cerebellum | 1 | 0.86 | (0.81,0.91) | 5.14E-08 |  | NA |
| *PSMG1\** | HFrEF | GTEx Stomach | 1 | 0.86 | (0.8,0.91) | 9.39E-07 |  | NA |
| *TAPT1* | HFrEF | GTEx Artery Aorta | 1 | 0.85 | (0.79,0.91) | 1.90E-06 |  | NA |
| *C6orf15* | HFrEF | GTEx Skin Not Sun Exposed Suprapubic | 1 | 0.85 | (0.79,0.9) | 1.89E-06 |  | NA |
| *MLIP\** | HFrEF | GTEx Artery Tibial | 1 | 0.81 | (0.75,0.88) | 1.08E-06 |  | NA |
| *SERPINF2* | HFrEF | deCODE | 6 | 0.80 | (0.75,0.86) | 4.98E-11 | -0.01; 0.66 | 0.72 |
| *DALRD3\** | HFrEF | GTEx Pancreas | 1 | 0.79 | (0.73,0.86) | 5.86E-08 |  | NA |
| *PABPC4\** | HFpEF | eQTLGen | 1 | 0.78 | (0.7,0.86) | 1.14E-06 |  | NA |
| *LIME1* | HFpEF | GTEx Muscle Skeletal | 1 | 0.77 | (0.69,0.85) | 9.84E-07 |  | NA |
| *FBXO32* | HFrEF | eQTLGen | 1 | 0.76 | (0.67,0.84) | 1.59E-06 |  | NA |
| *FBXL20\** | HFrEF | GTEx Artery Tibial | 1 | 0.75 | (0.67,0.84) | 6.16E-07 |  | NA |
| *NSF* | HFpEF | GTEx Esophagus Mucosa | 1 | 0.74 | (0.65,0.83) | 8.08E-07 |  | NA |
| *ROCK2* | HFrEF | GTEx Muscle Skeletal | 1 | 0.71 | (0.63,0.79) | 5.69E-10 |  | NA |
| *NDUFAF3* | HFrEF | GTEx Muscle Skeletal | 1 | 0.70 | (0.6,0.8) | 9.62E-07 |  | NA |
| *TCTA\** | HFrEF | GTEx Thyroid | 1 | 0.68 | (0.59,0.78) | 5.00E-08 |  | NA |
| *TNFRSF6B* | HFpEF | Fenland | 1 | 0.64 | (0.53,0.76) | 9.84E-07 |  | NA |
| *C3orf62\** | HFrEF | eQTLGen | 1 | 0.63 | (0.52,0.73) | 4.96E-08 |  | NA |
| *LTN1* | HFrEF | GTEx Muscle Skeletal | 1 | 0.58 | (0.49,0.68) | 6.21E-11 |  | NA |
| **Partially novel hits** |  |  |  |  |  |  |  |  |
| *ZBTB17* | HFrEF | eQTLGen | 1 | 2.56 | (1.85,3.27) | 3.81E-11 |  | NA |
| *PGAP3\** | HFrEF | GTEx Adipose Subcutaneous | 1 | 1.42 | (1.27,1.57) | 2.70E-11 |  | NA |
| *PHIP* | HFrEF | GTEx Thyroid | 1 | 1.34 | (1.18,1.5) | 2.06E-06 |  | NA |
| *IRAK1BP1\** | HFrEF | GTEx Adipose Visceral Omentum | 1 | 1.12 | (1.07,1.17) | 1.61E-06 |  | NA |
| *FUT11\** | HFrEF | GTEx Esophagus Mucosa | 1 | 1.10 | (1.06,1.14) | 8.68E-07 |  | NA |
| *MST1* | HFrEF | deCODE | 6 | 1.04 | (1.03,1.05) | 4.13E-12 | 0.02; 0.47 | 0.05 |
| *B3GNT8\** | HFrEF | deCODE | 4 | 0.96 | (0.94,0.97) | 6.91E-10 | -0.04; 0.17 | 0.59 |
| *MYOZ1* | HFrEF | GTEx Heart Atrial Appendage | 2 | 0.94 | (0.92,0.96) | 2.60E-12 |  | 0.94 |
| *TNFSF12\** | HFrEF | deCODE | 3 | 0.91 | (0.88,0.94) | 3.71E-08 | 0.01; 0.94 | 0.09 |
| *CELSR2\** | HFrEF | GTEx Muscle Skeletal | 1 | 0.89 | (0.85,0.93) | 1.04E-08 |  | NA |
| *MIA3* | HFrEF | GTEx Cells Cultured fibroblasts | 2 | 0.85 | (0.79,0.9) | 1.51E-06 |  | 0.69 |
| *PSRC1\** | HFrEF | GTEx Brain Cortex | 1 | 0.83 | (0.78,0.89) | 6.96E-09 |  | NA |
| *STRN* | HFrEF | GTEx Heart Atrial Appendage | 1 | 0.82 | (0.75,0.88) | 1.18E-06 |  | NA |
| *MAP3K7CL* | HFrEF | GTEx Cells Cultured fibroblasts | 1 | 0.77 | (0.71,0.83) | 8.42E-12 |  | NA |

### MR assumptions

Proposed instruments for our MR hits had a median (interquartile range, “IQR”) F-statistic of 76.08 (45.62, 177.32), suggesting weak instrument bias was unlikely to be observed (Figure 2). Multiple steps were conducted to assess confounding by linkage disequilibrium (LD). First, we performed genetic colocalization and observed a median (IQR) posterior probability of the shared causal variant hypothesis 4 (H4) of 0.85 (0.55, 0.93) (**Table S4**). Three HFpEF and 44 HFrEF genes had strong evidence of colocalization (H4 ≥ 0.8), while four HFpEF and 10 HFrEF genes had suggestive evidence of colocalization (0.5 ≤ H4 < 0.8). Second, we investigated if there was a HF, HFrEF, or HFpEF GWAS or MR-hit (including our hits) within +/− 500KB in LD, defined as r^2^ > 0.4, with the *cis*-QTLs of our MR hits (**Table S5**). Twenty-four MR hits were not in LD, eight hits were in moderate LD (0.4 < r^2^ < 0.8) and 48 MR hits were in strong LD (r^2^ > 0.8). A total of five HFpEF hits in chromosome 17 (*NSF*, *ARL17A*, *LRRC37A*, *LRRC37A2*, and *MAPT*) covering a region of 466 MB were in moderate LD. We used SuSiE in this region for fine-mapping in the presence of multiple causal variants, but the signals could not be distinguished due to the large LD block in the region. No fine-mapping was attempted for hits in strong LD given the limited utility of available resources. We did not observe heterogeneity (Cochran’s Q *p*-value > 0.05) across IV estimates in 11 of 15 MR hits with two or more instruments; in all five MR hits with three or more instruments, the Egger intercept test had a *p*-value > 0.05 (**Table 1**). In 39 of 66 (60%) hits that used *cis*-eQTLs as a proposed instrument, the Enformer CAGE value was ≥50% in tissues of the derived MR hit, confirming that a considerable number of our proposed instruments affected gene expression (**Table S6**).

**Figure 2.**
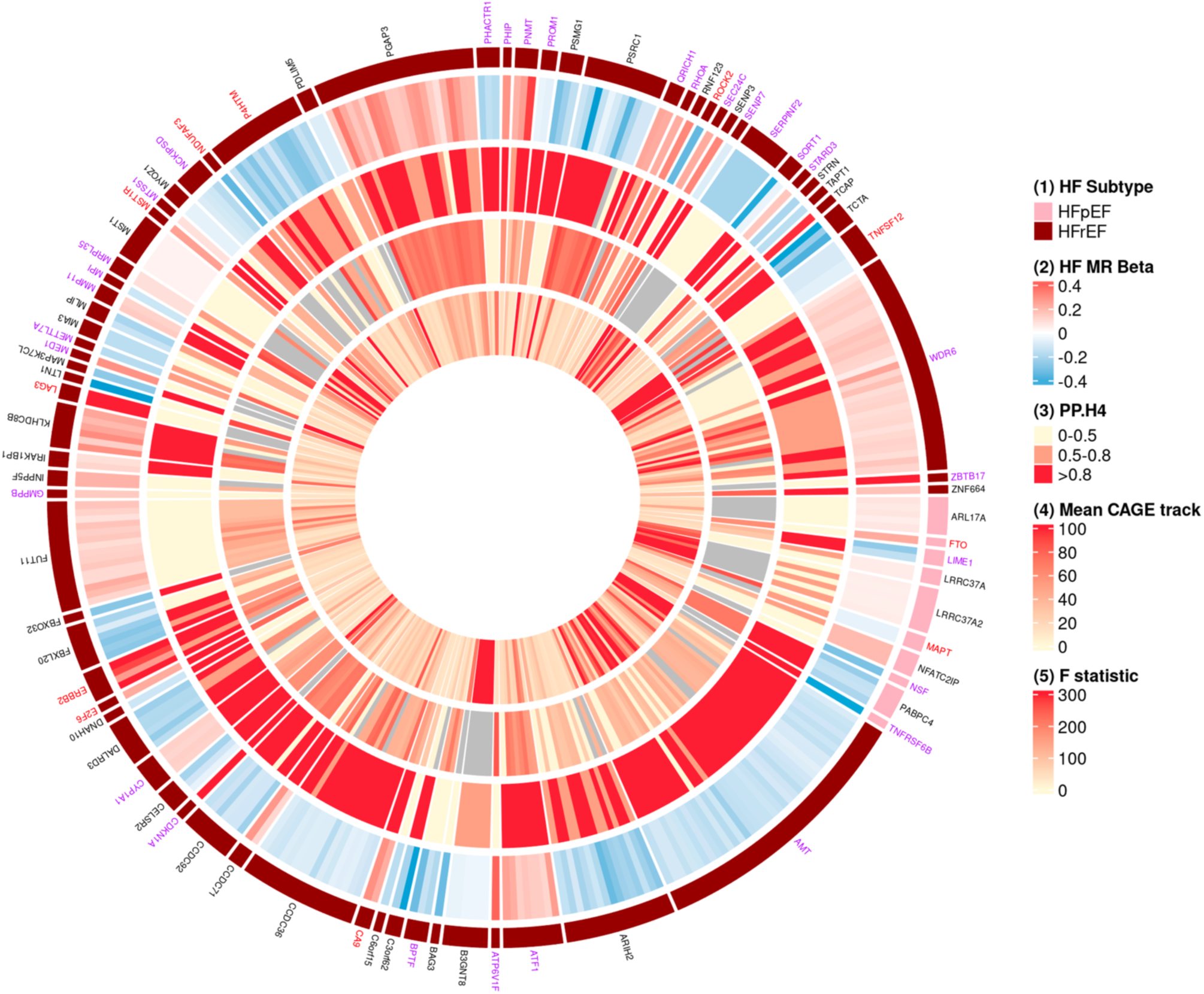
Robustness of MR hits against HFrEF and HFpEF. The length of the arc of the outer circle is proportional to the number of sources that the MR was significant for that gene. The gene names are colored according to druggability, based on information provided by Open Targets (release 2021-03-21). Gene names colored in purple correspond to potentially druggable, which includes targets of preclinical small molecules and of predicted druggable by small molecules or antibodies. Gene names colored in red correspond to targets of known drugs, which includes targets of approved drugs. Genes that do not fall in either of these two categories are colored in black. The outermost track (track #1) corresponds to the gene identified for HFrEF (in dark red) or HFpEF (in pink). The next track (track #2) going inward corresponds to the beta coefficient obtained in the MR analysis (colored from blue corresponding to a negative beta to red corresponding to a positive beta). The subsequent track (track #3) going inward corresponds to the colocalization PP.H4 (grouped by PP.H4 0-0.5, 0.5-0.8, and >0.8). The subsequent track (track #4) going inward corresponds to the mean CAGE value, presented for the tissue corresponding to the MR hit. The innermost track (track #5) corresponds to the variant F statistic.

### Therapeutic target profile

We then performed follow-up analyses for each hit to create a therapeutic target profile composed of supporting data on efficacy, novelty of biological mechanism, on-target safety, predicting MoA needed for a therapeutic solution, and druggability annotations. Orthogonal sources for efficacy included data from knockout (KO) mice models, putative loss of function (pLOF) variants identified at *p*-value < 5 × 10^−5^ from UK Biobank (UKBB), OMIM/ClinVar, and 29 cardiac MRI (CMR) parameters as proxies of structural heart disease. Sources utilized for safety included FDA warnings for drugs covering identified targets and MR results on 24 traits covering common causes of drug toxicity (cardiac, renal, and hepatic), as well as safety events typically included in phase 3 HF trials. The *p*-value threshold for follow-up analysis was 5 × 10^−5^ (0.05 / number of CMR, HF risk factors and safety traits tested).

### Orthogonal sources supporting efficacy on HFrEF and HFpEF

#### Structural Heart Disease

Seven of 10 HFpEF genes and 43 of 70 HFrEF genes had an association (*p*-value < 5 × 10^−5^) with at least one of the 29 evaluated CMR traits (**Table S7**). In 123 of 180 (68.3%) gene-CMR associations, the directionality of the MR on the CMR trait was concordant with the MR directionality on HFrEF or HFpEF. Interestingly, 21% of discordant associations were accounted for by LVMVR, which may be explained by uncertainty in the *directionality* of this metric with HF.^24^ We noticed that *ATP6V1F*, *KLHDC8B*, *ZBTB17*, *LTN1*, *C3orf62*, and *TCTA* showed strong associations with CMR traits, defined as greater than two standard deviations from the mean beta coefficient of the MR betas on CMR traits for the hits.

### Evidence from KO mice models, pLOF burden test in the UKBB, and OMIM/ClinVar on HF risk factors, HF, and cardiomyopathy-related traits

HFrEF hits *BAG3* and *TCAP* had orthogonal supporting evidence on HF or cardiomyopathy from KO mice models and OMIM/ClinVar, while *ARIH2*, *CCDC92*, *CDKN1A*, *ERBB2*, *INPP5F*, *MED1*, *MLIP*, *PDLIM5*, *PHIP*, *PNMT*, and *QRICH1*, all HFrEF hits, and *PABPC4*, a HFpEF hit, had orthogonal supporting evidence on HF or cardiomyopathy from only KO mice models (**Table S8**). An additional 13 gene-hits had orthogonal supporting evidence on HF risk factors from KO mice models or pLOF burden test (*p*-value < 5 × 10^−5^) from UKBB^25^ (Figure 3).

**Figure 3.**
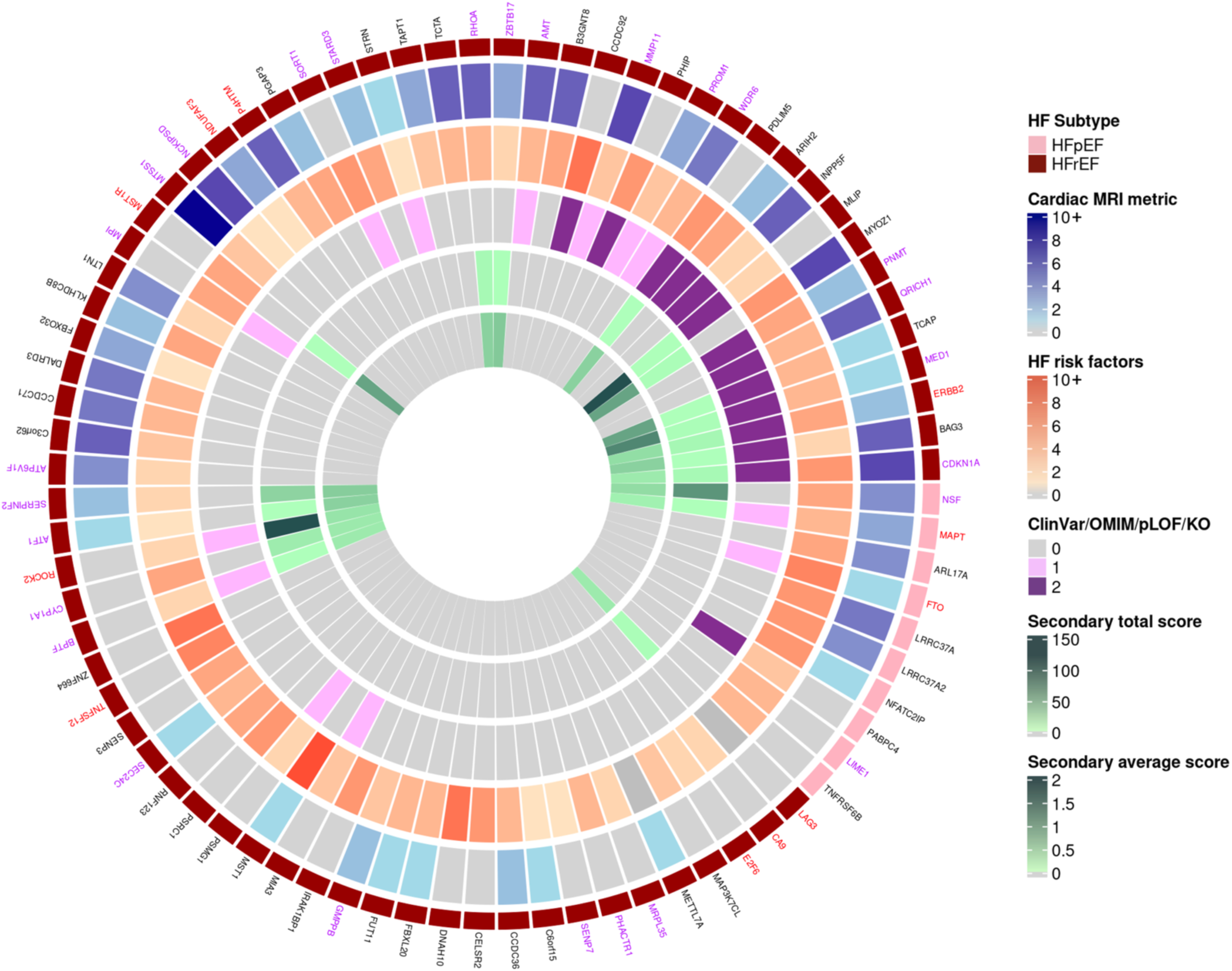
Orthogonal sources of support on efficacy. The gene names are colored according to druggability, based on information provided by Open Targets (release 2021-03-21). Gene names colored in purple correspond to potentially druggable, which includes targets of preclinical small molecules and of predicted druggable by small molecules or antibodies. Gene names colored in red correspond to targets of known drugs, which includes targets of approved drugs. Genes that do not fall in either of these two categories are colored in black. The outermost track corresponds to the gene identified for HFrEF (in dark red) or HFpEF (in pink). Following the outermost track, the next two tracks going inward correspond to the number of cardiac MRI metrics and the number of HF risk factors with *p*-value < 5 × 10^−5^ in the MR analysis for each gene. The next track going inward corresponds to results from ClinVar, OMIM, pLOF in UK Biobank, and knockout (KO) mouse models, where a “0” corresponds to no findings found for HF risk factors, a “1” corresponds to at least one finding for HF risk factor, and a “2” corresponds to at least one positive finding for HF/cardiomyopathy from any of the sources. The next two tracks going inward correspond to evidence from orthogonal sources of support on HF/cardiomyopathy obtained for secondary genes, where the “secondary total score” represents the total number of positive sources across all secondary genes that are linked to the same primary gene and “secondary average score” is determined by the ratio of the total score to the number of secondary genes associated with each primary gene.

### Supportive evidence from orthogonal sources of efficacy on secondary genes

Given that genes exhibit their downstream effects through biological networks, we identified all genes directly linked to our MR hits, referred to as secondary genes, using protein-protein interactions (PPI) networks, as described by MacNamara^26^. For every secondary gene, we retrieved data from KO mice models, pLOF burden test in the UKBB (*p*-value < 5 × 10^−5^), and OMIM/ClinVar on HF and cardiomyopathy-related traits and reported a score that considered the number of secondary genes linked to an MR hit. MR hits with the highest scores were *MLIP*, *MED1*, *MYOZ1*, *TCAP*, *MST1R*, *ZBTB17*, *SERPINF2*, *RHOA*, *HSPA4*, and *BAG3*, all HFrEF hits (Figure 3 and **Table S9**).

### Novelty of mechanism

Six HFpEF and 30 HFrEF genes had a novel biological mechanism defined as neither being the target nor being linked to a gene-target of HF drugs nor sharing a biological pathway with genes encoding targets for HF drugs. A closer inspection revealed that 31 of the 44 gene-hits judged as not novel were classified as such due to their link to the androgen receptor (AR), one of the two targets of spironolactone, which is unlikely to be the target explaining the benefits of this drug on HF^27^. The number of novel targets did not decrease after limiting the analysis to gene-hits for which >80% of their MR associations were with HF risk factors judged modifiable by existing cardiovascular drugs (coronary heart disease (CHD), LDL-C, blood pressure (BP), NT-proBNP, and type 2 diabetes mellitus (T2DM)) or by successful public health measures (smoking) (**Table S10**).

### Integration of human genetics sources to infer MoA for HFrEF and HFpEF hits

Except for *LAG3*, all hits had at least one MR association at a *p*-value < 5 × 10^−5^ with a CMR trait, HF risk factor, pLOF burden in UKBB, support by OMIM/ClinVar, or animal genetics. We then used this data to infer the MoA needed for efficacy by comparing the *directionality* (not expected to be affected by power) of findings on HFrEF or HFpEF against the orthogonal sources supporting efficacy. The concordance with HFrEF or HFpEF findings was 88% for HF risk factors (**Table S11**), 73% for CMR (73%), 50% (2 out of 4) pLOF burden test in the UKBB and OMIM/ClinVar, and 43% for KO mice models positive for HF (6 out of 14). The median (IQR) of the percentage of concordance, across gene-hits, between HFrEF and HFpEF findings with HF risk factors or CMR was 83% (67%, 100%), supporting MR as a valid strategy to infer MoA. Genes to highlight due to the high number of concordant associations with CMR and HF risk factors include the HFrEF hits *CDKN1A*, *BAG3*, *ZBTB17*, *MTSS1*, *MMP11*, *PDLIM5*, *MLIP*, *LTN1*, *CCDC92*, *MST1*, and *MST1R* and the HFpEF hits *FTO*, *NFATC2IP*, and genes in the chromosome 17 cluster (*NSF, ARL17A, LRRC37A*, and *LRRC37A2*) (**Table S12)**.

Given the unmet clinical need in HFpEF, we also report on CMR and HF risk factor associations for the 39 HFpEF suggestive hits (**Table S13**). Interestingly, the MR beta for HFpEF was highly correlated with the MR beta for BMI (*ρ* = 0.79) and T2DM (*ρ* = 0.89) across both Bonferroni and suggestive MR hits (Figure 4).

**Figure 4.**
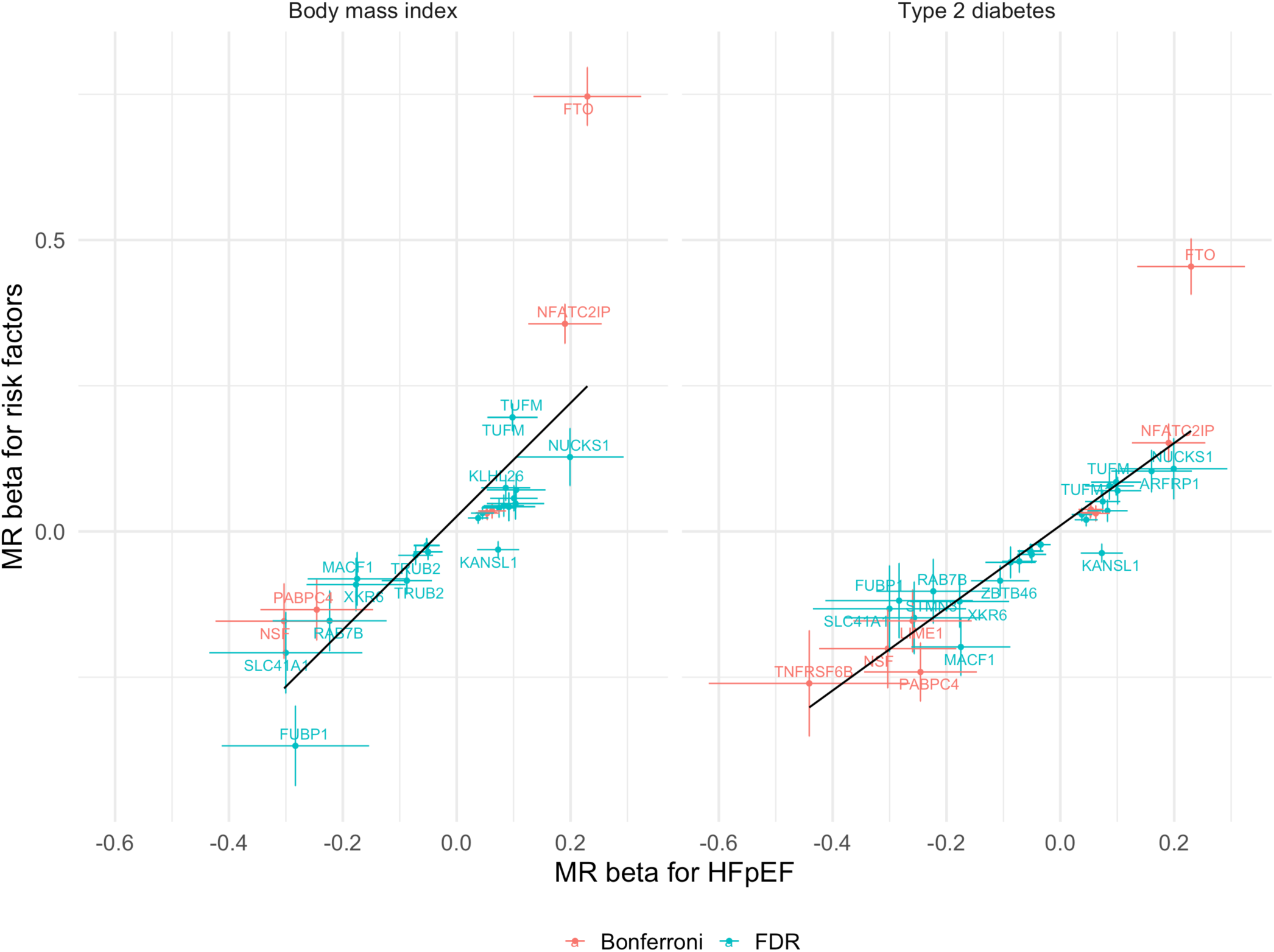
MR effect on HFpEF against the MR effect on HF risk factors, body mass index and type 2 diabetes, for HFpEF gene-hits. Plot is shown for HFpEF hits that pass the Bonferroni-adjusted threshold of *p*-value < 2.06 × 10^−6^ (in red) and for hits that pass the false discovery rate (FDR) of 5% (*p*-value < 6.80 × 10^−4^) (in blue). The error bars indicate the 95% confidence interval of the MR beta coefficient. The best-fit line derived through ordinary least squares is shown in black. The predicted model for body mass index is *y* = 0.03 + 0.97*x* (left plot) and for type 2 diabetes is *y* = 0.01 + 0.71*x* (right plot). The correlation between the MR betas for HFpEF and body mass index is 0.79 and between the MR betas for HFpEF and type 2 diabetes is 0.89.

### Safety

Of 1,920 gene-safety trait comparisons conducted, only 64 gene-safety traits (3.3%) had both a *p*-value < 5 × 10^−5^ and an MR beta coefficient direction of effect that was suggestive of a safety signal (Figure 5**; Table S14**). The most common safety signals were for liver (n=28), QT interval (n=21), renal (n=6) and cancer (n=8) traits. Interestingly, for 103 gene-safety trait comparisons, although they met a *p*-value < 5 × 10^−5^, the direction of the beta coefficient was indicative of a potential beneficial effect, instead of a safety event. In addition, we searched for FDA warnings reported for drugs whose only target was a protein encoded by our hits and identified tyrosine kinase inhibitors (e.g. anti-*ERBB2*^28^) and *LAG3* inhibitors that belong to the class of immune checkpoint inhibitors, both known for their cardiotoxicity^29^. None of our HFrEF and HFpEF hits were directly linked to the *KCNH2*/*hERG* gene associated with long QT prolongation syndrome^30^.

**Figure 5.**
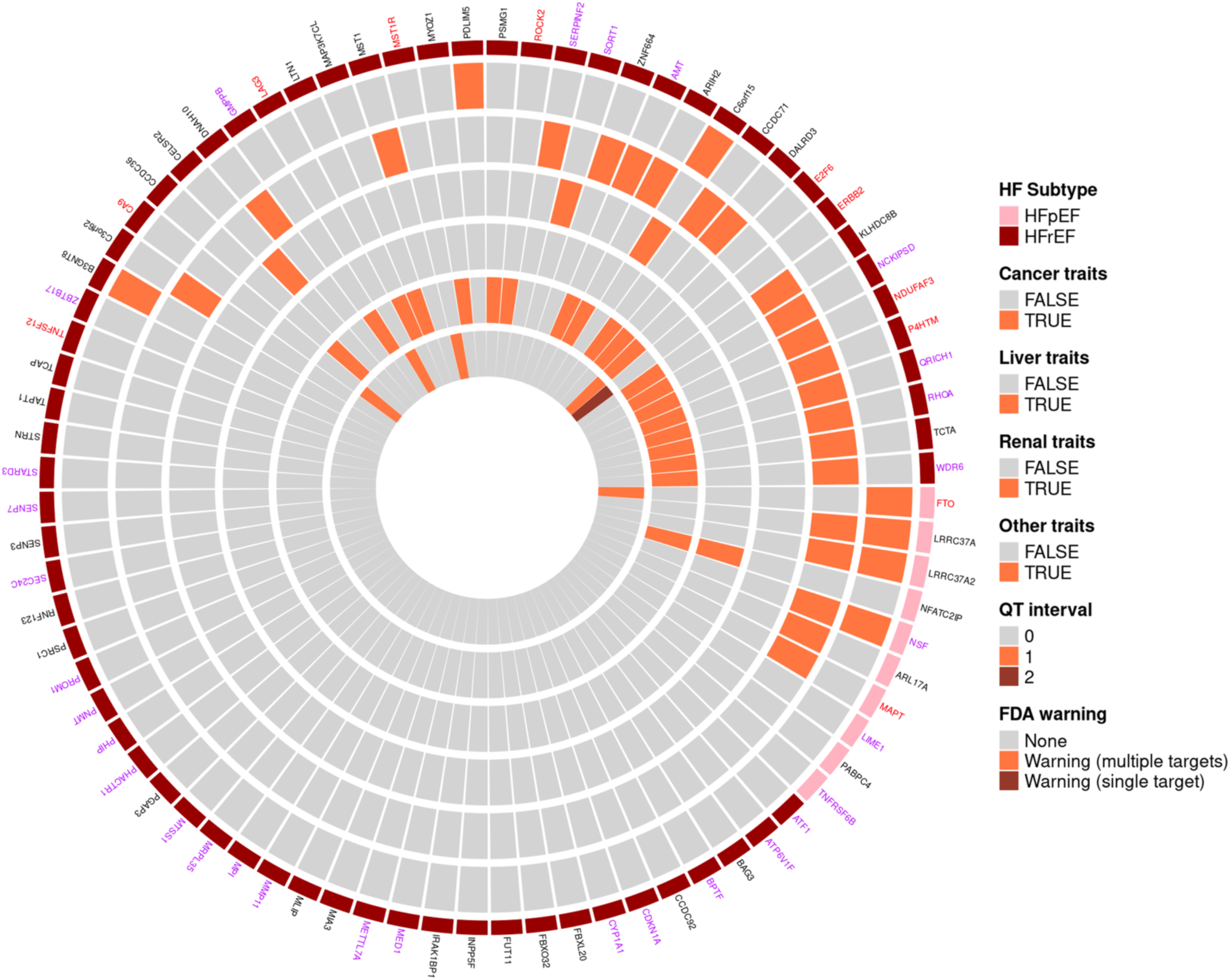
On-target safety for MR hits on HFrEF and HFpEF. The gene names are colored according to druggability, based on information provided by Open Targets (release 2021-03-21). Gene names colored in purple correspond to potentially druggable, which includes targets of preclinical small molecules and of predicted druggable by small molecules or antibodies. Gene names colored in red correspond to targets of known drugs, which includes targets of approved drugs. Genes that do not fall in either of these two categories are colored in black. The outermost track corresponds to the gene identified for HFrEF (in dark red) or HFpEF (in pink). Following the outermost track, the next four tracks going inward correspond to on-target safety traits, classified by cancer (second track going inward), liver (third track), renal (fourth track), and other traits (fifth track). Cancer traits include breast cancer, colorectal cancer, lung cancer, and prostate cancer. Liver traits include serum alanine aminotransferase (ALT), alkaline phosphatase, aspartate aminotransferase (AST), chronic liver disease and cirrhosis, and gamma-glutamyl transferase (GGT). Renal traits include creatinine, cystatin C, proteinuria, renal failure, acute renal failure, chronic renal failure (CKD), and trefoil factor 3. Other traits include pulmonary heart disease, atrioventricular (AV) block, paroxysmal ventricular tachycardia, acute pulmonary heart disease, chronic pulmonary heart disease, Alzheimer’s disease, and total creatine kinase. The next track (sixth track) corresponds to QT interval (“0” indicates that the gene is not linked to the *HERG* gene for QT syndrome and there is no QT interval as a potential safety trait, “1” indicates QT interval is identified as a potential safety trait, and “2” indicates the gene is linked to the *HERG* gene). The next track (seventh track) going inward corresponds to FDA warning, where “None” corresponds to no drug and no FDA warning, and “Warning (single targets)” and “Warning (multiple target)” corresponds to a reported FDA warning for a drug with single target or multiple target, respectively.

### Druggability

Twenty-seven HFrEF hits and five HFpEF hits were considered potentially druggable by small molecules or antibodies^31–33^, according to Open Targets (release 2021-03-08). We identified 30 drugs where the HFrEF hits *ROCK2*, *TNFSF12*, *CA9*, *ERBB2*, *LAG3*, and *MST1R* and HFpEF hits *MAPT* and *FTO* were the only efficacy target and hence have some translational opportunities. Of these 30 drugs, only Narnatumab, a neutralizing monoclonal antibody that blocks the MST1 ligand from binding *MST1R* (both MR hits), had a MoA that matched the MR findings on HFrEF for *MST1* and *MST1R* and had no reports on cardiotoxicity^34^. Narnatumab was discontinued after a Phase 1 trial in patients with advanced solid tumors, as it did not observe changes in pharmacodynamic biomarkers and antitumor activity (**Table S15**).

### Rediscovery of drugs approved or under development for HF management

At nominal *p*-value, we rediscover genes encoding targets of HF approved drugs, such as *ADRB1* for beta-blockers, *SCNN1A* for diuretics, *NR3C2* for mineralocorticoid receptor antagonist, *CACNA1D* for calcium channel blockers, and *SLC5A2* for *SGLT2* inhibitor (**Table S16**). As suggestive hits (*p*-value < 6.80 × 10^−4^) for HFrEF, we identified *LPA*, *IL6R*, *ADM,* and *EDNRA,* all associated with investigational drugs for cardiovascular disorders. Specifically, *LPA* is the target of several liver-targeted RNA-based therapies (coinciding with the liver GTEx tissue source of the MR) in development for cardiovascular disorders^35^. We further investigated *LPA* using 67 variants derived from a GWAS on Lp(a) from UKBB^36^. We replicated the association with HFrEF (*p*-value = 6.89 × 10^−9^), and showed a suggestive association with HFpEF (*p*-value = 1.67 × 10^−4^) (**Table S17**). Similar associations with HFrEF and HFpEF were observed when using MR-pQTL from Olink UKBB (**Table S18**). Based on the 94% reduction in Lp(a) levels observed with 75 mg olpasiran^37^, the OR on HFrEF is expected to decrease by 25% and on HFpEF by 15%. *IL6R*, whose ligand (IL6) is the target of Ziltivekimab, is being tested for progression of HFmEF (heart failure with mid-range ejection fraction) and HFpEF in the HERMES Phase 3 trial^38^. To confirm these findings, we used 22 cis-*IL6R* variants associated with CRP levels as proxy of IL6R blockade and showed a stronger association with HFrEF (*p*-value < 3.41 × 10^−9^) but a null effect on HFpEF (*p*-value = 0.47) (**Table S19**). *ADM,* target of Enibarcimab, is being tested for cardiogenic shock. Although there are several dual endothelin receptor antagonists, our findings on HFrEF with *EDNRA* suggest selective ET_A_ antagonists as a potential target for HF management.

### Replication of findings

After conducting a multi-ancestry meta-analysis on African-American, Hispanic, and European-descent individuals from the BioVU and MVP, including 12,604 HFrEF and 11,486 HFpEF cases that were not included in the discovery phase (**Table S20**), we conducted MR using *cis*-QTLs utilized in the MVP European-descent discovery dataset. A total of 31 HFrEF hits and 4 HFpEF hits had a *p*-value < 0.05, and of these, 94.3% were also directionally concordant with the discovery dataset (**Table S21**). As an additional replication, we used Olink Explore 3072 assay from the UK Biobank. We conducted MR using 1,682 *cis*-pQTL, covering the same number of genes, against HFrEF and HFpEF. A total of seven genes passed the Bonferroni correction threshold (*p*-value < 2.97 × 10^−5^), of which six were a replication (*ZBTB17*, *CELSR2*, *TNFRSF6B*, *LRRC372A2*, and *MST1* for HFrEF and *LPA* for HFrEF and HFpEF) and one, *MUC2*, was a new discovery for HFpEF (details in **Table S18**).

## 3. DISCUSSION

Our analysis explored the causal roles of the human proteome and transcriptome on the largest HFrEF and HFpEF genetics dataset and identified 58 novel targets^8,9,18–23^, for which we developed a therapeutic target profile encompassing efficacy, safety, and novelty of mechanism to help inform early drug discovery programs for HFrEF and HFpEF.

The novelty of our findings comes from multiple avenues. First, we accounted for pathophysiologic differences between HFrEF and HFpEF to explore genetic signals reported as hits for unclassified HF and, in some cases, strengthen the supporting evidence as HF drug targets. Second, we identified several genes, discussed below, as potential targets for HFrEF or HFpEF with low likelihood of confounding of genetic evidence by LD, significant support from orthogonal sources on efficacy, compelling evidence around the MoA needed for therapeutic efficacy, and no evidence for an unacceptable safety profile. Third, we identified several biological pathways as potential therapeutic targets for HF management.

We report new evidence relevant to drug discovery for *MTSS1* and *ZBTB17*, previously reported hits for HF. For *MTSS1*, we refined the association to HFrEF. Second, using different resources compared to previous reports (GTEx instead of MAGNet and CMR instead of echocardiography)^39^, we showed that the *cis*-variant rs7461129/*MTSS1* which is associated with increased mRNA expression in LV tissue was associated with increased risk in HFrEF. In addition, we report ten associations with LV and RV MRI parameters, nine of which are directionally concordant with HFrEF findings, suggesting an inhibitor as a therapeutic agent. This MoA is also supported by animal genetics where ablation of *MTSS1* gene showed a beneficial HF profile^39^.

Our previous MVP GWAS on HF subtypes identified rs371236917 as a hit for HFrEF; however, both *ZBTB17* and *HSPB7* genes were considered candidate genes. Using eQTLGen and subsequently confirmed by Olink (5×10^−14^, with same directionality), we now show that *ZBTB17* had the strongest MR association with HFrEF and a substantial increase in LVESV and reduction in LVEF. These findings suggest an inhibitor as the MoA for a therapeutic solution, which agrees with in-vitro, neonatal rat cardiomyocytes, and in-vivo mouse models where overexpression of *ZBTB17* led to cardiac hypertrophy^40^. In-vitro experiments have also suggested that *ZBTB17* binds to both calcineurin and *NFATC2* known factors in both cardiac remodeling and HF; these are mechanisms that could explain associations with HF observed in human and animal genetics^40^.

Among the nine novel genes for HFpEF, it is important to highlight the *NFATC2* interacting protein (*NFATC2IP*) and poly(A) binding protein cytoplasmic 4 (*PABPC4*) genes. MR findings on *NFATC2IP* showed both an increase in risk on HFpEF and higher LVM. Increase in HFpEF risk was also concordant with increased levels of systolic BP, CRP, and BMI, increased risk of T2DM, renal failure, and pulmonary heart disease, and a decrease in HDL-C levels—traits highly relevant for HFpEF. rs3768321/*PABPC4* variant, which increases mRNA expression of *PABPC4* in multiple tissues showed a reduction in HFpEF risk. This result was directionally concordant with MR findings showing a reduction in levels of BMI, risk of T2DM, and renal failure, and an increase of HDL-C levels. In agreement with the MR findings, genetic ablation of *PABPC4* in mice showed a reduction in HDL-C and free fatty acids and an increase in heart weight^41^. The concordance in the directionality of the findings between human and animal genetics studies supports an agonist for HFpEF prevention. Further support for the role of *PABPC4* on HFpEF comes from GWAS studies, where our *cis*-instrument rs3768321/*PABPC4* has been reported as a hit for metabolic traits such as basal metabolic rate, whole body fat free mass, FEV1, and peak-expiratory flow, reinforcing the role of *PABPC4* on cardiometabolic fitness^31^.

Among our novel HFrEF gene-hits that are important to highlight are *PDZ,* LIM domain 5 (*PDLIM5*), and muscular LMNA interacting protein (*MLIP*). *PDLIM5* is a cytoskeleton-related protein, highly expressed in heart tissue, that interacts with multiple sarcomeric components, protein kinases, and transcription factors and is involved in cell proliferation and cardiomyocyte physiology^42^. Our MR findings showed that an increase in mRNA expression of *PDLIM5* exhibited a protective effect against HFrEF and cardiomyopathy in MVP participants. These findings were directionally concordant with a reduction in diastolic BP and an improvement in renal function, proxied by reductions in creatinine and cystatin C levels and an increase in eGFR levels. In support of this, global and cardiac specific ablation of *PDLIM5* gene in mice resulted in dilated cardiomyopathy phenotype, all of which indicates that agonism of *PDLIM5* is the MoA for a HFrEF therapy ^43^.

*MLIP* gene, highly expressed in heart tissue, interacts with *LMNA*, an established cardiomyopathy gene. MLIP protein appears to regulate cardiac homeostasis and protect against cardiac hypertrophy. In agreement with these mechanistic findings, we observed that a *cis*-variant associated with an increase in mRNA expression of *MLIP* showed a protective effect on HFrEF and cardiomyopathy in MVP. In support of this, *MLIP* overexpression in mice models abrogates adverse cardiac remodeling and preserves cardiac function, while ablation of *MLIP* showed a rapid progression from hypertrophy to HF^44^.

It did not escape our attention that genes we identified together with those previously reported converged on pathways such as ubiquitin-proteasome system (*UPS*), small ubiquitin-related modifier (SUMO) pathway, inflammation, and mitochondrial metabolism known to play a role on HF etiology.

Compensatory mechanisms relevant to HF, such as LV hypertrophy, are associated with both an increase in protein synthesis and degradation. Impairments to UPS, essential in protein degradation, may lead to an increase in abnormal proteins, which is detrimental to the heart given its limited self-renewal capacity^45^. *PSMG1*, *RNF123*, *LTN1*, *ARIH2*, *FBXL20*, and *FBXO32* gene-hits and *FBXO31* and *WWP1* suggestive gene-hits all belong to the UPS pathway. All of these genes were associated with HFrEF. Evidence from protein-coding variants supporting the role of UPS in HF comes from LOF variants in the *FBXL4* gene, known to cause mitochondrial DNA depletion syndrome, which includes hypertrophic cardiomyopathy within its phenotypic expression^46^. In support of these human genetic findings on UPS genes, large-scale KO-mice studies from the International Mouse Phenotyping Consortia identified additional F-box proteins (FBX10, 15, 22, 24, 36, 38) as hits for measures of structural heart disorders using cardiac ultrasound^47^. Evidence from human and animal genetics adds to the growing mechanistic evidence from preclinical and clinical studies suggesting a role for UPS in HF^48^. However, recent cardiotoxicity findings associated with proteasome inhibitors (targeting 38 proteins that compose the 26S-proteasome) licensed for management of plasma cell dyscrasias^49^, suggests that determining the specific target of intervention within UPS, MoA, and dosing are critical gaps to be surmounted to achieve beneficial effects on HFrEF^50^.

SUMO system is composed of two processes, SUMOylation and deSUMOylation, involved in the regulation of cardiac development, metabolism, and stress adaptation, all central mechanisms to HF^50,51^. SUMOylation is a process by which SUMO proteins (SUMO1, 2, 3, and 4) are conjugated to target proteins through a pathway controlled by the enzymes, E1 activating, E2 conjugation, and E3 ligases. Overexpression in mice of *SUMO1* and *2* was associated with HF phenotypes^51^, while high plasma levels of *SUMO3* have been reported in prevalent HF in the AGES Reykjavik study^52^ and SUMO2 is associated with HF progression^53^. In support of this, our analysis identified *UBA6*, *UBA7* (E1 activating enzymes), and *UBE2E3* (E2 conjugation enzyme) genes as suggestive hits for HFrEF. DeSUMOylation is the reverse process by which SUMO-specific peptidases (SENP) remove SUMOs from protein targets. Our analysis identified *SENP3* and *SENP7* as hits for HFrEF, while mice models overexpressing *SENP2* and *SENP5* were associated with cardiac hypertrophy and dilated cardiomyopathy^51^. Hence, our data suggests the potential benefit of targeting the SUMO pathway for benefit in HFrEF.

After the post-hoc analysis of the CANTOS trial, which evaluated the IL1β inhibitor canakinumab, showed a trend towards dose-dependent reduction in hospitalization for HF or HF-related mortality^54^, efforts for novel anti-inflammatory targets in HF have focused on the NOD-like receptor pyrin domain-containing protein 3, cryopyrin (*NLRP3*)/*IL1*/*IL6* signaling pathway^38,55,56^. From upstream of the pathway, we identified the coiled-coil domain containing 92 (*CCDC92*) gene, encoding an interferon stimulated protein, as a hit for HFrEF. Genetic ablation of *CCDC92* in mice, after a high fat diet, is associated with a reduction in *NLRP3* and IL1β serum levels and metabolic traits such as obesity and insulin resistance^57^. Associations of *CCDC92* gene with metabolic and inflammatory traits have also been observed by GWAS studies, using the missense variant rs11057401/*CCDC92*, and by our MR downstream analysis using *cis*-eQTLs from LV and adipose tissues in strong LD (r^2^=0.9) with rs11057401^58^. IL1 receptor associated kinase-1 binding protein (*IRAK1BP1*) gene, identified by us as a hit for HFrEF, is a component of the IRAK1 molecule involved in IL1R proximal signaling. Further down the pathway, the *IL6R* gene, reported as a MR hit for unclassified HF,^9,18^ was identified by us as a suggestive hit for HFrEF, but not for HFpEF. The specificity of the association of *IL6R* with HFrEF (*p*-value < 3.41 × 10^−9^), instead of with HFpEF (*p*-value = 0.47), was strengthened when using 22 *cis*-*IL6R* variants associated with CRP levels, as a proxy of downstream *IL6R* blockade.^59^ The *IL6R* findings specific to HFrEF contrast with the target population being evaluated in the HERMES trial (HFmEF/HFpEF) testing a mAb-targeting *IL6*^38^. Post-*IL6R*, several signaling molecules (such as IL6ST, STAT1 and STAT3, and TRAF3) have been reported as hits by GWAS or MR studies for unclassified HF^9,10,18^. Though the human genetic evidence for the NLRP3-IL1-IL6 signaling pathway on HF is compelling, the evidence from general population biobanks emulates a primary prevention trial on HF, as opposed to trials on HF progression. Hence, further human genetic studies that use biobanks such as MVP, which emulate HF progression trials, are needed.

A growing number of clinical studies that evaluated energetics in HF patients have pointed towards a mitochondrial dysfunction affecting patients with HFrEF and HFpEF^60,61^. In support of this, inherited mitochondrial disorders, caused by a growing list of rare variants in mitochondrial-related genes, are known to cause cardiomyopathy, in the absence of established HF risk factors, as part of its multi-morbidity^60^. However, whether alterations in those genes through *cis*-variants affecting expression or cognate protein levels are relevant for common forms of HF has remained elusive. Our current study identified three genes involved in mitochondrial translation and assembly. *MRPL35* and *NDUFAF3* (hits) and *MRPS21* (suggestive) are associated with HFrEF and structural measures of heart disease in our study. This provides further support to the consensus statement that has identified mitochondrial function as a therapeutic target for HF^61^.

Strengths from the current analysis include the large number of HFrEF and HFpEF cases in a uniformly phenotyped cohort allied with multiple orthogonal approaches to establish causal relations of druggable protein targets. We undertook extensive analytical steps to reduce the likelihood of confounding by LD, and our analyses have a low chance of weak instrument bias, as reinforced by the finding that six out of ten of the MR hits used *cis*-instruments, judged by Enformer to be true positive variants affecting *cis*-transcription in the same tissue used by MR. Given that some of the MR assumptions are unverifiable^62^, we triangulate our MR findings with orthogonal sources, vulnerable to different biases, to prioritize detected MR hits. Our emphasis on concordance of the directionality of findings between HFrEF/HFpEF, HF risk factors, and CMR allowed us in several cases to infer MoA for targets identified with confidence. We conducted multiple steps to increase the replicability of our findings to other studies and ancestries using BioVU and MVP and the Olink platform.

There are important limitations to our study. A considerable number of our MR hits were in strong LD with themselves and with published hits from GWAS/MR studies on HF, HFrEF, HFpEF, or cardiomyopathy genes. In situations with strong LD (r^2^>0.8), fine-mapping methods have low value to resolve this. Hence, we triangulated with orthogonal sources, exposed to different biases; however, this was uninformative in multiple cases. De-novo experimental studies are needed to resolve confounding by strong LD observed in some cases and to provide some biological insight for hits with low biological prior evidence, but we considered this to be out of scope for our current analysis. In addition, no single cell model (e.g. iPSC cardiomyocyte) will prove suitable for all our hits. Furthermore, for several hits we deemed it to be unlikely that such experimental evidence will drastically change the likelihood of hits being relevant to HF given the existing evidence we identified from orthogonal sources. Although we included non-European ancestry samples in our replication datasets, the eQTL/pQTL instruments for MR were not ancestry-specific and assumed transportability across ethnicities. This highlights the need for resources such as GTEx to include non-European individuals.

In conclusion, we identified several genes as plausible targets for HFrEF and HFpEF and found evidence from orthogonal sources to support their efficacy and inform on the MoA needed for a therapeutic solution. We anticipate this knowledge to be of value in the generation of novel therapeutic opportunities for HFrEF and HFpEF.

## 4. METHODS

### 4.1 Clinical and demographic characteristics of HF genomic datasets

The discovery dataset consisted of 422,645 participants of European-descent (27,799 patients with HFrEF, 27,579 with HFpEF, and 367,267 control individuals) included in MVP Release 4. The clinical and demographic features of the participants are summarized in **Table S20**. Replication was conducted across ancestries in MVP Release 4, using data on 108,202 participants of African American descent (7,393 patients with HFrEF, 6,515 with HFpEF, and 94,294 controls) and 46,622 participants of Hispanic descent (1,865 patients with HFrEF, 1,904 with HFpEF, and 42,853 control individuals, **Table S20**). Replication was also conducted across studies in the Vanderbilt University Medical Center (VUMC) Biobank (BioVU) using data on 17,744 participants of European descent (2,807 with HFrEF, 2,610 with HFpEF, and 12,327 control individuals) and 2,640 participants of African-American descent (539 with HFrEF, 457 with HFpEF, and 1,644 control individuals, **Table S20**).

#### 4.1.1 Million Veteran Program

The MVP is a national voluntary research program and mega-biobank in the health care system of the Department of Veterans Affairs (VA). MVP contains comprehensive data linking genotype data to electronic health record (EHR) data, containing information on diagnosis codes, laboratory values, and imaging reports. Enrollment in the MVP began in 2011 with veterans recruited from over 60 medical centers. Data was collected from participants with available VA EHR data. All participants have provided informed consent, and the study protocol of the MVP was approved by the Veterans Affairs Central Institutional Review Board. The study design and methodology of the MVP has been previously described^7^.

Participants with HF in MVP were identified with an International Classification of Diseases (ICD)-9 code of 428.x or ICD-10 code of 150.x and with an echocardiogram performed within 6 months of diagnosis. Participants with heart failure were then classified into the subtypes of heart failure, HFrEF and HFpEF, by implementing a natural language processing (NLP) approach on both coded and unstructured data in the VA EHR in order to extract left ventricular ejection fraction (LVEF) values from echocardiogram reports^63^. Participants with an HF diagnostic code were classified as HFpEF if their first recorded LVEF was greater than or equal to 50% and as HFrEF if their first recorded LVEF was less than or equal to 40%. Participants were identified as having European, African American, or Hispanic ancestry using a combination of self-reported race and ethnicity and principal components of ancestry using previously described methods^64^.

#### 4.1.2 Genotyping and quality control in the Million Veteran Program

Genotyping in MVP was performed using the MVP 1.0 custom Axiom array, which is based on the Axiom Genotyping Platform. The MVP 1.0 custom Axiom array was designed and developed as a single assay to be used on clinically useful and rare variants applicable to the multi-ethnic MVP cohort^65^. The Axiom array consists of 668,280 genetic markers that pass quality control. Quality control involved removing duplicated, mislabeled, and misidentified samples, samples with excess relatedness (identified by a call rate below 98.5%), and samples with gender mismatches^65^. Imputation was then conducted on the 1000 Genomes phase 3 version 5 reference data using Minimac4^66^. Variants with a minor allele frequency (MAF) greater than 1% were tested for association with HFpEF and HFrEF using PLINK2.^67^ Covariates included age, sex, and the top ten genotype-derived principal components. GWAS findings were functionally annotated and prioritized using FUMA, a software that uses information from 18 biological data repositories and tools^68^.

#### 4.1.3 Vanderbilt University Medical Center’s BioVU

The BioVU is a biobank that links the de-identified EHR system containing phenotypic data to discarded blood samples from routine clinical testing for the extraction of genetic data^69^. Participants with HFrEF and HFpEF were identified by the Electronic Medical Records and Genomics (eMERGE) network phenotype definition for incident HFrEF and HFpEF, which includes electronic medical record (EMR) International Classification of Diseases, Ninth and Tenth Revision (ICD-9 and ICD-10) codes (ICD-9 code 428.x and ICD-10 code I50.x) and clinical notes. The algorithm uses ICD-9 and ICD-10 codes, problem list mentions, and HF mentions from clinical text identified by natural language processing. The EMR-driven phenotype algorithm of heart failure was conducted according to a previously published method^70^, which includes three hierarchical definitions of heart failure (definite, probable, and possible). Only individuals who have been flagged as “definite” and “probable” were included; individuals who have been flagged as “possible” were excluded from analyses. Echocardiography measurements of left ventricular ejection fraction were extracted from a structured database^70^. NLP was used to search radiology reports for EF measurements^70^, where a classification of HFpEF required the individual’s lowest ejection fraction to be greater than or equal to 50%, and HFrEF required the individual’s lowest ejection fraction to be less than 40%. HF cases with an intermediate ejection fraction (greater than or equal to 40% or less than 50%) were not included in the analyses. Individuals whose medical records lacked any ICD codes for heart failure or text mentions and who never had a documented left ventricular ejection fraction below or equal to 50% in their echocardiogram reports were used as controls. Age was defined as age at the time of heart failure diagnosis for cases and age at last medical visit for controls.

##### 4.1.3.1 Genotyping and quality control in the BioVU

GWAS-level genotyping was performed using the Illumina MEGA-Ex chip, which includes over 2 million common and rare variants prior to imputation^69^. Quality control involved excluding samples or variants with greater than or equal to 5% missingness, removing samples with mismatched identifiers as detected by checks for identity by descent, and removing samples that were discordant between reported sex and genetically determined sex^69^.

### 4.2 Mendelian randomization on 15,253 genes and HFrEF and HFpEF in MVP

We performed MR analysis using eQTLs and pQTLs spanning 15,253 unique protein-coding genes across 5 different data sources as instrumental variables to infer effects of protein and expression levels on HFrEF and HFpEF.

#### 4.2.1 Selection of proposed eQTL and pQTL instruments

We obtained *cis*-pQTLs from publicly available GWAS from the Fenland (retrieved from www.omicscience.org), deCODE, and ARIC studies, all using SOMAscan v4 assay in plasma and from European-ancestry individuals^12–14^ (**Figure S2**). The deCODE study provided genetic association data on plasma protein levels of 4,907 proteins measured in 35,559 individuals, Fenland provided data on 3,892 proteins measured in 10,708 individuals, while ARIC provided data on 2,004 proteins measured in 7,213 individuals. To diminish the likelihood of horizontal pleiotropy, instruments were restricted to *cis*-variants. In the Fenland study, approximate conditional analysis was performed to detect secondary signals for each genomic region identified by distance-based clumping of association statistics^8^. In the Fenland study, we used a total of 2,900 *cis*-pQTLs across 1,557 genes covering an equal number of proteins that passed a Bonferroni threshold of *p*-value < 1 × 10^−11^. In the ARIC study, we selected *cis*-pQTLs that were identified as significant independent variants from the original study, which included a total of 2,004 significant SOMAmers that had at least one *cis*-pQTL (that met an FDR threshold of 5%) near the gene of the putative protein. We used unconditional estimates from this list of 2,004 *cis*-pQTLs from the original ARIC study. *Cis*-variants in both deCODE and ARIC were selected based on conditional analysis. A total of 2,004 cis-pQTLs covering an equal number of proteins from the ARIC study were used as proposed instrumental variables. From the deCODE study, we selected *cis*-pQTLs that were identified as significant (*p*-value < 1 × 10^−7^) independent variants from the study, and removed duplicates by chromosome and position, resulting in 5,662 instruments across 1,663 genes encoding a protein (1,674 proteins and 1,703 SeqIDs). In the ARIC study, *cis*-regions were defined as +/− 500kb of the transcription start site (TSS), whereas in the Fenland and deCODE studies, *cis*-variants were defined as +/− 1Mb of the transcription start site (TSS) of a protein-coding gene. We note that despite independent *cis*-variants were identified through conditional analysis by the study authors in Fenland, deCODE, and ARIC, we used the unconditional beta coefficients to avoid bias by using beta coefficients derived from conditional analysis. Across all sources, we have used *cis*-variants with unconditional beta coefficients.

For protein-coding genes that are not covered by pQTLs, we used summary statistics derived from expression quantitative trait locus (eQTL) for whole blood from 31,684 individuals from the eQTLGen Consortium, and for an additional 48 tissues, we derived eQTLs from Genotype Tissue Expression Project (GTEx) Version 8 using raw data^16^. We utilized data across 48 tissues of GTEx because we leveraged eQTLGen data for whole blood. For GTEx, we selected independent *cis*-eQTLs per gene by performing up to five conditional analyses in regions (+/− 1 Mb from the transcription start side of each gene) using individual-level data in GTEx Version 8, adjusting for the peak variant if the association passed a *p*-value of 1 × 10^−4^. The primary signal is unconditional. To identify independent signals, we considered primary and conditional associations that passed a *p*-value of 5 × 10^−8^ and extracted the effect size and standard error estimates from the unconditional association. For eQTLGen, we defined instruments using the lowest *p*-value per gene in the publicly available summary statistics files. We computed the beta and SE for these files using the reported Z-scores (*Z*) and *p*-values (*p*) using the formula 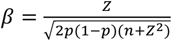 and 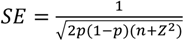.

Genes with instruments covered by multiple pQTL sources were all used; genes with instruments covered by multiple eQTL sources (and which did not have data from any pQTL source) were all used. If the proposed IV variants were not present in the outcome GWAS, then a proxy variant was selected based on a *r*^2^ greater than or equal to 0.8 between the two variants using the 1000 Genomes Project European reference panel. Palindromic variants and non-protein-coding genes were removed.

#### 4.2.2 Two-sample Mendelian randomization

The effects of eQTLs and pQTLs on HFrEF and HFpEF were assessed using the inverse variance weighted method for genes with two or more IVs and the Wald ratio method for genes with a single IV using TwoSampleMR version 0.5.6 (release date 03-25-2021)^71^. TwoSampleMR was conducted across each dataset, including Fenland, deCODE, ARIC, eQTLGen, and for each of the 48 tissues in GTEx. We defined MR hits as those passing our Bonferroni correction criteria (*p*-value < 2.06 × 10^−6^), which was determined by dividing 0.05 by the number of tests conducted (n = 24,257), which is the total number of unique genes in each QTL source, including 1,383 genes in Fenland, 1,342 in ARIC, 1,493 in deCODE, 8,054 in eQTLGen, and 11,985 in GTEx. If a gene passed this threshold in more than one QTL source (including tissues for eQTLs or datasets for pQTLs), then its beta coefficient must be directionally concordant across all such sources. Because Bonferroni is a conservative threshold, we also identified suggestive hits using a Benjamini-Hochberg false discovery rate (FDR) of 5%, corresponding to a *p*-value < 6.80 × 10^−4^, which is more lenient than our Bonferroni threshold.

To define novelty of hits, we compared our findings against all previously published GWAS and MR studies in HF and HF subtypes^8,9,18–23^ and categorized hits as one of three categories: novel, partially novel, and replication. Novel genes are defined as those that have not been previously reported and are not in LD (*r*^2^ < 0.4) with a variant that has been published as a GWAS hit or as an instrument used for a MR hit; for further details, see section on “assessment of confounding by LD”. Partially novel genes are defined as genes that have been previously associated with unclassified HF, but that our study was able to assign to HFrEF or HFpEF. Otherwise, the MR hit was considered a replication of previous studies.

### 4.3 Follow up analysis on MR hits

For all genes that passed a MR Bonferroni corrected *p*-value < 2.06 × 10^−6^, we assessed MR limitations and established a therapeutic target profile including efficacy, on-target safety, novelty of mechanism of action, and druggability opportunities.

#### 4.3.1 MR Sensitivity Analyses

##### 4.3.1.1 Confounding due to Linkage disequilibrium

To assess the likelihood of confounding due to LD, we performed genetic colocalization analysis of eQTLs and pQTLs with HF subtypes, examined the 1 Mb region around each proposed IV from each MR hit, and searched for variants previously reported by GWAS and MR studies on HF and HF subtypes. First, we performed genetic colocalization to evaluate whether our *cis*-pQTLs and eQTLs shared at least a single causal variant with HFpEF and HFrEF. We assigned a prior of 1 × 10^−4^ for *p*_1_ and *p*_2_, the prior probability that a variant in the region is associated with trait 1 and trait 2, respectively, and 5 × 10^−5^ for *p*_12_, the prior probability that a variant is associated with both traits. To limit bias from pleiotropy, we used a colocalization window around a lead variant of +/− 250kb. A posterior probability of hypothesis 4 (PP.H4) greater than 0.8 was considered as strong evidence for colocalization at the variant level, and PP.H4 greater than 0.5 but lower than 0.8 was considered suggestive evidence of colocalization at the variant level. All colocalization analyses were performed using the coloc R package (version 5.2.2)^72^. We also examined the 1 Mb region (+/− 500 kb) around each proposed instrument to identify any GWAS variants and MR genes that have been identified in nine previously published studies for unclassified HF, HFrEF, HFpEF as well as MR hits from the current analysis within the region. For instruments with a HF or HF subtype hit from any of nine previously published GWAS or MR studies as well as our own within a +/− 500 kb region, we used the 1000 Genomes Project European reference panel to measure linkage disequilibrium by calculating the *r*^2^ between our proposed variant and the identified nearby variant. We considered an *r*^2^ greater than 0.8 as high LD and between 0.4 to 0.8 as moderate LD, see **Table S5**. We also searched for known cardiomyopathy genes within a 1MB region around the MR hits.

###### Fine-mapping using SuSiE for determining causal variants for MR hits that are in LD with previously known HF loci

We used the R package MungeSumstats to harmonize the two datasets with the reference genome GRCh37 and dbSNP^73^. INDELs were dropped from this analysis. LD was extracted from the GTEx v8 data. The packages coloc:SuSiE^74^ were used to identify credible causal sets. SuSiE^75,76^ (“Sum of Single Effects”) is a fine-mapping method based on a new model for sparse multiple regression, and entails a Bayesian modification of simple forward selection. The model outputs several credible sets with high probability for containing a variable with non-zero effect. The region chr17:42972122-44500000, as evident from the regional plot, contained a large LD block; therefore, the identified fine-mapped set were very large (containing 1,372 out of the 1,920 SNPs in the same set), making fine-mapping very difficult to conduct in this region. We did not perform SuSiE for hits identified in high LD (*r*^2^ ≥ 0.8), given the limited utility of SuSiE for performing fine-mapping in these settings.

##### 4.3.1.2 Weak instrument bias and horizontal pleiotropy

To assess weak instrument bias, we computed two key parameters from the first-stage regression of the exposure phenotype on the genetic variant: the proportion of variance explained (*R*^2^) and the F-statistic. We calculated *R*^2^ as a function of effect size estimate *β*, the minor allele frequency MAF, the standard error of effect size (se), and the sample size *n*^77^:

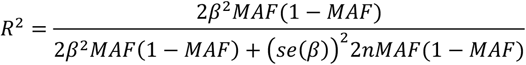

We computed F-statistic^78^ using the formula 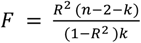, for the number of instrumental variables, *k*, and the sample size, *n*. We used a threshold of F < 10 to denote a weak instrumental variable^79^. To assess the compatibility of the IV estimates, we conducted a test of heterogeneity and report the heterogeneity *p*-value, where a *p*-value below 0.05 was considered heterogeneous. To evaluate directional pleiotropy, we performed the MR-Egger intercept test for genes with three or more instruments. Directional pleiotropy was considered present when the MR-Egger intercept differed from the null (*P* < 0.05).

##### 4.3.1.3 Further validation of *cis*-eQTL instruments for MR hits

For all MR hits that used *cis*-eQTLs as instruments, we used Enformer to evaluate if these *cis*-eQTLs are likely to be causal in gene expression. Enformer aims to predict gene expression from DNA sequence with a deep learning approach^80^. The output is composed of 5,313 genomic tracks for the human genome and 1,643 genomic tracks for the mouse genome. The four types of predicted genome-wide tracks (CAGE, ChIP Histone, ChIP TF, Dnase/ATAC) concern various cell types and tissues. We directed our attention to the 638 human genome CAGE tracks. We start with an input of 257,642 rsID-GTEx-tissue triplets used as MR instruments across 49 GTEx tissues. After combining with Gencode information, this is reduced to 257,613. A further 16,749 variants were excluded because they were further away than 57,344 base pairs from the transcription start site (TSS), as defined by GTEx GENCODE 26 (gencode.v26.GRCh38.genes.txt file retrieved from **https://storage.googleapis.com/gtex_analysis_v8/reference/gencode.v26.GRCh38.genes.gtf**). A total of 240,819 variants-GTEx-tissue triplets covering 23,304 unique genes remained in the Enformer analysis as described below. For each CAGE track, we assigned a percentile value for each variant-gene pair (i.e. we obtain a matrix of 638 values per variant-gene pairs). Briefly, we annotated each variant-gene-tissue with evidence from a causal model, with the mean percentile annotated across 638 CAGE outputs from Enformer. The percentile indicates where the statistics for a specific gene-variant of a specific CAGE track falls compared to the distribution of the entire data range. The percentile values are calculated on the distribution of the statistic over the CAGE track for all the variants.

The mean CAGE track value across all CAGE tracks is used to summarize the overall effect of a SNP on gene expression. We considered a mean CAGE track value greater than or equal to 50 to be supportive evidence for a particular *cis*-eQTL to be causal on its cognate gene expression. This threshold was chosen because 50 is the expected mean value given that the mean CAGE track values are uniformly distributed. Additionally, when we examined the entire SNP-gene dataset, particularly the bottom 250 variants with the lowest F-statistics, 50 was the observed average, further supporting the use of this threshold in the analysis. We noticed that by ranking all the SNPs by their F-statistics, the top SNPs had significantly higher Enformer percentiles (*p*-value = 2.7 × 10^−8^) for the Kolmogorov-Smirnov test comparing SNPs with top and bottom F-statistics and mean CAGE score, providing support for the instruments used.

##### 4.3.1.4 Colocalization across pQTL sources

We used colocalization analysis to assess shared genetic etiology across the 3 pQTL sources (deCODE, ARIC, and Fenland). Specifically, for genes with a significant MR result, we performed colocalization between all pairs of *cis*-variants available from the QTL sources passing MR. We used the same coloc parameters and thresholds as that in the “Confounding due to Linkage disequilibrium” section. All analyses were conducted using the coloc package in R.

##### 4.3.1.5 GWAS Meta-analysis for replication

To replicate our findings, we performed a GWAS meta-analysis on 175,208 individuals, including 12,604 HFrEF and 11,486 HFpEF cases of European and African-American descent from the BioVU, and 9,258 HFrEF and 8,419 HFpEF cases of African-American and Hispanic descent from the MVP. The BioVU replication cohort consisted of 2,807 HFrEF and 2,610 HFpEF cases of European-descent and 539 HFrEF and 457 HFpEF cases of African-American descent. The MVP replication cohort consisted of 7,393 HFrEF and 6,515 HFpEF cases of African-American descent and 1,865 HFrEF cases and 1,904 HFpEF cases of Hispanic descent. We performed the GWAS meta-analysis using a fixed-effects model, and we used the inverse-variance-weighted (IVW) average method, which summarizes the effect size from multiple independent studies by calculating the weighted mean of the effect sizes using the inverse variance of the individual studies as weights. Allele frequency was calculated as the averaged frequency across studies. All analyses were conducted using the METAL software (version released 2020-10-06)^81^.

##### 4.3.1.6 Olink validation

We performed a validation study using Olink Explore 3072 data generated by the UK Biobank Pharma Proteomics Project^82^ (UKB-PPP) characterizing plasma proteomic profiles of 52,363 participants from the UK Biobank. The Olink platform consists of 2,941 immunoassays targeting 2,925 unique proteins. The Olink Explore 3072 proximity extension assay (PEA) platform is based on a pair of polyclonal antibodies.^83^ The antibodies in the Olink platform attach to different locations on the target protein and are marked with single-stranded complementary oligonucleotides. When pairs of antibodies that match bind to the protein, the linked oligonucleotides hybridize, and are subsequently detected through next-generation sequencing^84^. The UK Biobank plasma samples were taken at Olink’s facilities in Uppsala Sweden and processed using NovaSeq 6000 Sequencing Systems. Further details of the Olink proteomics assay, data processing, and quality control have been previously described^82^. Consistent with our main analysis, instruments were restricted to *cis*-variants (defined as within 1 Mb from the gene encoding the protein) and we used unconditional beta coefficients to run two-sample MR. We used a total of 2,057 *cis*-pQTLs covering an equal amount of genes that were identified as significant (*p*-value < 1.7 × 10^−11^) independent variants from the study. We removed palindromic variants and variants that were not in our outcome dataset, resulting in a final total of 1,682 *cis*-pQTLs that were utilized as instrumental variables. The pQTL mapping was conducted on the full UK Biobank PPP cohort on 52,363 participants and up to 23.8 million imputed variants. We conducted two-sample MR on the harmonized datasets to estimate effects on HFpEF and HFrEF consistent with methods detailed in the main analysis.

##### 4.3.1.7 Lipoprotein(a) Mendelian randomization analysis on HFpEF and HFrEF

We performed two-sample MR on lipoprotein(a) [Lp(a)] concentration on HFpEF and HFrEF using genetic association effect estimates from Lp(a) variants from a genome-wide association study on Lp(a) on 293,274 participants of the UK Biobank^36^. Lp(a) (nmol/L) was measured using an immunoturbidimetric analysis on a Randox AU5800, and measurements that returned an error or were outside the reportable range for Lp(a) (3.80-189 nmol/L) were excluded from analyses^36^. Associations of genetic variants with natural log-transformed Lp(a) were tested in linear regression models, controlling for age, sex, genotype batch, and 20 principal components of ancestry. We performed clumping on variants that passed genome-wide significance (*p*-value < 5 × 10^−8^) using the PLINK clumping method and a European reference panel, where variants in LD within a window of 1 Mb and with an r^2^ < 0.01 were pruned. The variant with the lowest *p*-value was retained. We removed palindromic variants and variants that did not harmonize with our outcome datasets. Using 67 variants, we performed inverse-variance weighted (IVW) MR on HFrEF and HFpEF. To estimate the causal effect adjusted for any directional pleiotropy, we performed MR-Egger. We computed linkage disequilibrium between these 67 variants and the *LPA* variant in GTEx liver (rs2292334) and Olink (rs56393506) by calculating r^2^ using the 1000 Genomes European reference panel.

A randomized, double-blind trial of olpasiran therapy at 34 participating sites across seven countries identified a significant reduction in Lp(a) concentrations in patients with established atherosclerotic cardiovascular disease^37^. The baseline median laboratory value of Lp(a) concentration (nmol/L) of trial participants who were given 75 mg olpasiran every 12 weeks (n = 58) was 227.5 (IQR 188.4-304.2). The percent change in the Lp(a) concentration from baseline to week 36 was −93.8 (95% CI −97.3 to −90.3). Therefore, we estimated the effect on HFrEF and HFpEF risk by multiplying the log of the ratio of endpoint to baseline Lp(a) concentration with 75 mg olpasiran (−2.786) by the MR beta, which represents the change in HFrEF or HFpEF per change in natural log-transformed Lp(a) (nmol/L). We exponentiated this value to convert to the percent reduction in the odds ratio of HFrEF or HFpEF.

#### 4.3.2 Therapeutic target profile

For every MR hit, we leveraged orthogonal sources of evidence and created a therapeutic target profile covering efficacy, on-target safety, novelty of mechanism and druggability opportunities. Orthogonal sources for efficacy included information on 29 cardiac MRI metrics used as proxies for structural heart damage, HF-related traits from putative loss-of-function (pLOF) in UK Biobank (UKBB), OMIM/ClinVar, animal models (knockout and transgenic), and evidence (OMIM/ClinVar, pLOF UKBB, and animal studies) from genes directly linked to the MR hits (referred to as “secondary genes”), identified through protein-protein interactions (PPI). Sources utilized for safety include FDA warnings for drugs covering identified targets, and MR results on 24 traits covering cardiac, renal, and hepatic toxicity, and safety events usually included in phase 3 HF trials. A *p*-value < 5 × 10^−5^ was used to denote statistical significance in follow-up analysis, which is less than 0.05/number of total tests conducted.

##### 4.3.2.1 Cardiac MRI traits

For every MR HF subtype hit, we determined the MR estimates for the following cardiac MRI metrics: left atrial (LA) emptying fraction, LA emptying volume, LA maximum volume, LA minimum volume, left ventricular (LV) end-diastolic volume, LV ejection fraction (LVEF), LV end-systolic volume(LVESV), LV mass (LVM), LV mass to volume ratio (LVMVR), LV stroke volume (LVSV), right ventricle (RV) end-diastolic volume (RVEDV), RV ejection fraction (RVEF), RV end-systolic volume (RVESV), and RV stroke volume (RVSV). For this, we used unpublished data from UKBB using data from 32,994 individuals^24^. We additionally determined MR estimates for myocardial interstitial fibrosis using publicly available GWAS^85^ and for left atrial volume, longitudinal peak diastolic strain rate, and radial peak diastolic strain rate using a publicly available GWAS^86^.

###### HF-related traits from pLOF burden test in UKBB, OMIM/ClinVar and animal models

To determine if our findings have been reported at both the phenotype and gene level, we examined multiple orthogonal sources of support, including variants of clinical significance from ClinVar^87,88^, Online Mendelian Inheritance in Man (OMIM)^87^, putative loss-of-function data from GeneBass^89^, and knockout models from Mouse Genome Informatics (MGI)^90^. We retrieved from ClinVar, a centralized repository of information about genomic variants and their clinical significance, data on loss-of-function variants, missense variants, and pathogenetic variants.^87,88^ We queried OMIM, a comprehensive catalog of human genes related to genetic disorders and traits^87^, for entries on any genetic disorders or traits linked to mutations or variations in the gene. Using the Genebass (gene-biobank association summary statistics) browser with UK Biobank data, we explored gene-based phenome-wide association study (PheWAS) analysis results to identify rare-variant association results^89^. Group tests were conducted using the mixed-model framework SAIGE-GENE, which includes single-variant tests and gene-based burden (mean) test^89^. We queried the Mouse Genome Informatics (MGI, http://informatics.jax.org/) resource and retrieved all cardiovascular system phenotypes attributed to mutations/alleles of the gene. Each entry for each of these orthogonal sources of support (ClinVar, OMIM, GeneBass, and MGI) have been meticulously reviewed by two independent reviewers to classify them as: (1) HF risk factor phenotype, and (2) HF and cardiomyopathy phenotype. We developed a summary score representing the overall number of positive sources identified per gene for both the HF risk factor phenotype and the HF and cardiomyopathy phenotype, where a positive finding in ClinVar or OMIM, GeneBass, and MGI were considered in the score.

###### Evidence from secondary genes

Given that genes, and hence drug targets, exhibit downstream effects through biological networks, we searched for genes directly linked to our MR-hits (referred as “secondary genes”) and retrieved information on HF-related traits from pLOF from UKBB, OMIM/ClinVar, and animal models (knockout and transgenic). Secondary genes were identified through a procedure introduced by MacNamara^26^ that aggregates various interaction networks to create a comprehensive protein-protein interactions (PPI) network. Resources used included STRING (score >= 500)^91^, the Complex Portal^92^, the Human Reference Interactome Mapping Project (Lit-BM)^93^, the OmniPath^94^, the Human Integrated Protein-Protein Interaction reference (HIPPE; score >= 0.9)^95–98^, and the Ligand-receptor interactions MetaBase. MetaBase contains over 6 million experimental findings on protein-protein, protein-nucleic acids, and protein-compound interactions. Lit-BM contains a human ‘all-by-all’ reference interactome map of human binary protein interactions, known as ‘HuRI’, containing over 53,000 protein-protein interactions.^93^ To harmonize the different resources, we converted different representations of gene IDs to gene symbols and for every gene we aggregated interactions across all resources. After harmonizing these PPI resources, the aggregated network contained 50,163 nodes and 13,750,500 interactions. Using this network, the secondary genes are defined as genes directly interact with the MR hits. Using data on orthogonal support on the secondary genes, we computed an average score for the level of orthogonal support across all secondary genes linked to a primary gene, which was computed as the total number of positive findings for ClinVar or OMIM, GeneBass, and MGI across all secondary genes divided by the number of secondary genes associated with each primary gene.

###### HF risk factors

We conducted two-sample MR, as described for HF subtypes, of our gene-hits for the following 20 HF risk factor traits: atrial fibrillation, atrial flutter, body mass index, chronic airway obstruction, troponin, diastolic blood pressure, estimated glomerular filtration rate, HDL-c, ischemic heart disease, LDL-c, systolic blood pressure, cardiac troponin I measurement, type 2 diabetes, NT-proBNP measurement, cardiomyopathy, primary and intrinsic cardiomyopathies, troponin I cardiac muscle, alcohol intake frequency, C-reactive protein, and current tobacco smoking. For current tobacco smoking, C-reactive protein, alcohol intake frequency, and troponin I cardiac muscle, we used publicly available GWAS summary statistics^99–101^, and for all other risk factors, we used data from a meta-analysis of three data sources, FinnGen, UK Biobank, and MVP^102^.

We conducted MR using pQTLs and eQTLs as instrumental variables on HF risk factors and safety traits using data from the MVP, UK Biobank^103^, and FinnGen version 7^104^. To harmonize phenotypes between the three sources, disease-based traits were mapped using phecodes and ICD-10 codes from the sources. UK Biobank traits with an ICD-10 code were mapped to phecodes; if the phecode had not already been mapped to MVP in the previous step, then a match was made, if possible. For all unmapped MVP phecodes with case counts lower than 4000, only MVP data was used for the analyses. To map MVP and UK Biobank phenotypes to FinnGen, NLP was used to assign phenotypes described in the three sources to the closest available Experimental Factor Ontology (EFO)^105^ codes. All matches were submitted to clinical adjudication. We considered findings significant at a *p*-value threshold < 5 × 10^−5^.

###### Novelty of Mechanism of Action (MoA)

We used two hard criteria and one soft criteria for identifying novel MoA for our hits. For the first hard criteria, we searched if our hits were the target, or directly linked to a target, of a drug approved for HF management. We retrieved information on HF drugs and its targets from ChemBL, followed by manual curation by two authors (JJ and JPC). To identify if our MR-hits were directly linked to gene targets of drugs used for HF management, we used our interaction network as described above.

For the second hard criteria, we investigated if our hits shared the same biological pathway of drug targets approved for HF management. To achieve this, we extracted biological pathways of every gene by querying the EnrichR database. This process extracts pathways with adjusted *p*-value < 0.05. Then, for every MR hit gene, we intersected its corresponding pathways with those associated with the drug-target genes.

For the soft criteria, we used two sample MR to determine the profile of HF risk factors associated with our hits. We considered hits that were exclusively or largely (>80%) associated with modifiable HF risk factors to be not novel and vice versa, hits that were exclusively or largely associated with no/partially modifiable HF risk factors or no HF risk factor to be novel. Modifiable HF risk factors were CHD, blood pressure, LDL-C, NT-proBNP, T2DM, and smoking. Non or partially modifiable HF risk factors used were HDL-C, eGFR, atrial fibrillation/ atrial flutter, alcohol intake frequency, body mass index, C-reactive protein (CRP) and COPD. CRP as a proxy (non-causal) of inflammation due to the *NLRP3/IL1B/IL6* pathway was considered a non-modifiable HF risk factor.

###### MR for on-target safety traits relevant to HF

We conducted two-sample MR, as described for HF subtypes, of our gene-hits against the following 24 safety traits: serum alanine aminotransferase (ALT) measurement, alkaline phosphatase, aspartate aminotransferase (AST) measurement, breast cancer, chronic liver disease and cirrhosis, colorectal cancer, creatinine, cystatin C, gamma glutamyltransferase (GGT), lung cancer, prostate cancer, proteinuria, pulmonary heart disease, atrioventricular block, paroxysmal ventricular tachycardia, renal failure, acute renal failure, chronic renal failure (CKD), acute pulmonary heart disease, chronic pulmonary heart disease, Alzheimer’s disease, total creatine kinase, trefoil factor 3, and QT interval. For trefoil factor 3 and QT interval, we used publicly available GWAS summary statistics^100,106^. For the remaining safety traits, we used data from a meta-analysis of 3 data sources, FinnGen, UK Biobank, and MVP, consistent with the approach utilized for the analysis of HF risk factors^102^.To be considered as a potential on-target safety signal, the MR on a safety trait must have reached a *p*-value threshold < 5 × 10^−5^ and the directionality of its beta coefficient must be discordant with that observed for HFrEF or HFpEF.

###### FDA box warnings

For gene-hits that are targets of drugs in clinical phase or licensed, we retrieved any FDA box warning associated with such drugs from Open Targets. To determine if a box warning is applicable to our MR hits, first, the drug must have only one drug-target and the MR beta on HF subtype must be concordant with the pharmacological action of the drug. A positive MR beta on HF subtype is concordant with drug inhibitors or negative modulators, while a negative MR beta is concordant with drug agonists or positive modulators. Information on pharmacological action for drugs was retrieved from ChemBL. The exception to this rule are drugs whose pharmacological action was described as “other”. To determine if our HF subtypes findings are the rediscovery of a box warning cardiotoxicity of existing drugs, the MR beta on HF subtype needs to be discordant with the pharmacological action of the drug and ideally the drug should have only one target; otherwise this was considered as a potential rediscovery as the cardiotoxicity could be due to other drug targets, for further details see **Figure S3.**

###### Druggability annotations

Protein-encoding genes identified as hits in our MR analyses for HF subtypes were annotated with drug tractability information based on information provided by OpenTargets^31–33^ (release date 2021-03-21), as described previously by our group^8^. For easier interpretation, we regrouped the original Open Targets buckets into four mutually exclusive groups: Category 1, which includes targets of approved drugs; Category 2, targets of drugs in clinical trials; Category 3, targets of preclinical small molecules; and Category 4, targets predicted druggable by small molecules or antibodies. The remaining proteins were considered non-druggable. For genes that were determined to be druggable, we extracted information on those drugs from ChEMBL v32^107^. For each drug with available information, we assigned all indications, clinical phase, and the mechanism of action (MoA) for the drug. To retrieve any possible gene-drug pairs that may have been missing from the Open Targets pipeline, we extracted absent Ensembl IDs for any proteins found in ChEMBL using Uniprot’s rest API.

For genes that were the target of licensed drugs, we checked whether the disease indication was also a risk factor for HF, as this may introduce a bias analogous to confounding by indication in MR. Translational opportunities were defined as those gene-hits that are the only target of a drug or compound and whose MR beta coefficient on HF subtype is concordant with the MoA of the drug. We also identified the targets of drugs used in HF management and determined how many of these targets were directly (or indirectly through secondary genes) rediscovered by our MR-QTL strategy. MR findings on drug-targets being pursued for HF, identified through clinical trials.gov, are reported regardless of *p*-value.

## Data Availability

All data produced in the present study are available upon reasonable request to the authors.

## Acknowledgements

We are grateful to all the MVP investigators; a list of MVP investigators can be found in the Supplementary Materials. This research is based on data from the Million Veteran Program, Office of Research and Development, Veterans Health Administration, and was supported by awards #MVP037 [BLR&D Merit Award BX005831], Veterans Affairs Grant I01CX001922 (PI: Joseph), and #MVP001 [I01-BX004821]. This publication does not represent the views of the Department of Veterans Affairs or the United States Government. We also acknowledge the VA Merit Grant I01-CX001025 (PI: Wilson/Cho).

The British Heart Foundation funded the manual analysis to create a cardiovascular magnetic resonance imaging reference standard for the UK Biobank imaging resource in 5000 CMR scans (www.bhf.org.uk; PG/14/89/31194). This work acknowledges the support of the National Institute for Health and Care Research Barts Biomedical Research Centre (NIHR203330); a delivery partnership of Barts Health NHS Trust, Queen Mary University of London, St George’s University Hospitals NHS Foundation Trust and St George’s University of London. This article is supported by the London Medical Imaging and Artificial Intelligence Centre for Value Based Healthcare (AI4VBH), which is funded from the Data to Early Diagnosis and Precision Medicine strand of the government’s Industrial Strategy Challenge Fund, managed and delivered by Innovate UK on behalf of UK Research and Innovation (UKRI). Views expressed are those of the authors and not necessarily those of the AI4VBH Consortium members, the NHS, Innovate UK, or UKRI. This work was funded by UKRI Programme Grant MC_UU_00002/18. For the purpose of open access, the author has applied a Creative Commons Attribution (CC BY) licence to any Author Accepted Manuscript version arising. SEP acknowledges support from the “SmartHeart” EPSRC programme grant (www.nihr.ac.uk; EP/P001009/1). SEP provides consultancy to Circle Cardiovascular Imaging, Inc., Calgary, Alberta, Canada. JW holds membership of scientific advisory boards/consultancy for Relation Therapeutics and Silence Therapeutics and ownership of GSK shares. NA acknowledges support from Medical Research Council for his Clinician Scientist Fellowship (MR/X020924/1). During the completion of this work, JPC became an employee of Novartis.

