## Supplementary Figures for "Large-scale Mendelian randomization identifies novel pathways as therapeutic targets for heart failure with reduced ejection fraction and with preserved ejection fraction"

**Figure S1. The number of unique and shared genes among the present study categorized as HFpEF/HFrEF MR, and prior studies on HF GWAS, HFpEF/HFrEF GWAS, HF MR, and cardiomyopathy presented as a Venn diagram (Figure S1A) and as an upset plot sorted by cardinality (Figure S1B).** In Figure S1A, the percentage presented in parentheses represents the proportion of the total data set that falls within each section of the Venn diagram. Sets with no genes are not annotated.

**(A)**

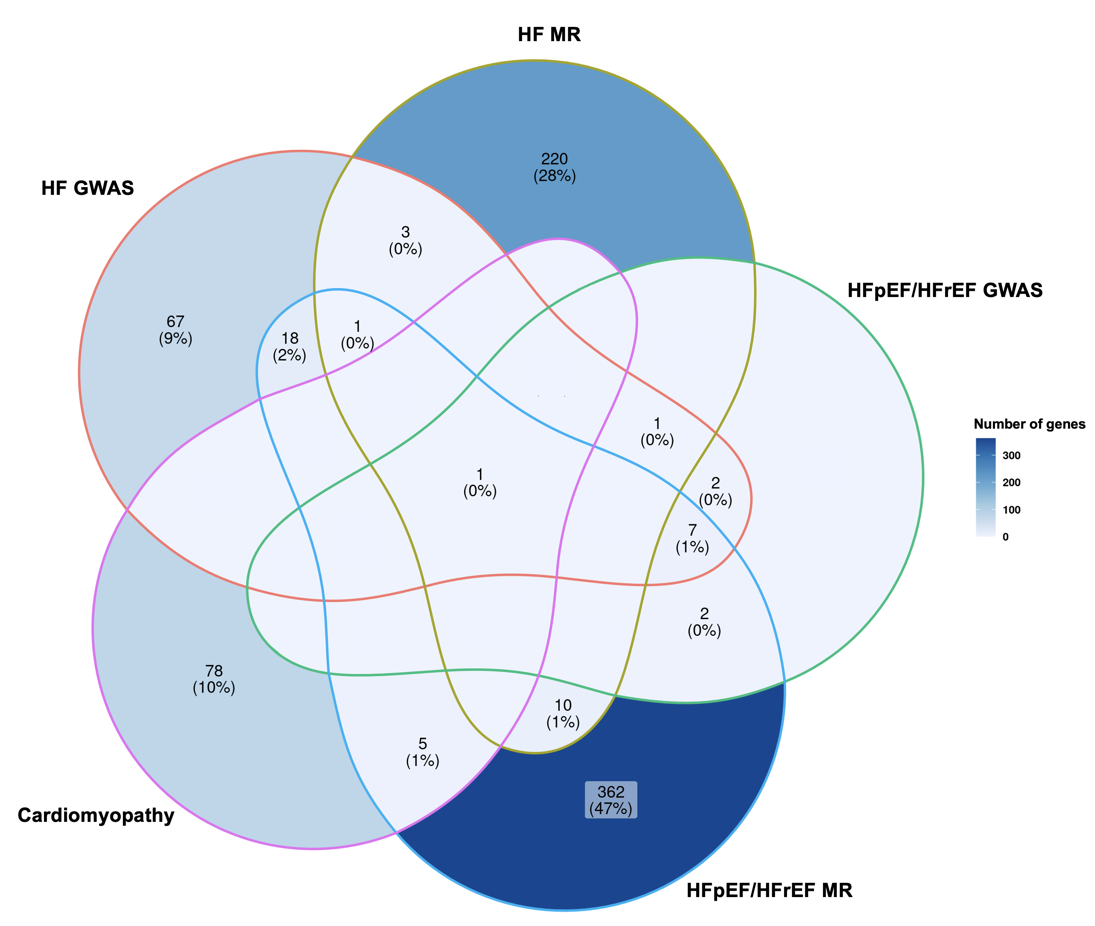

**(B)**

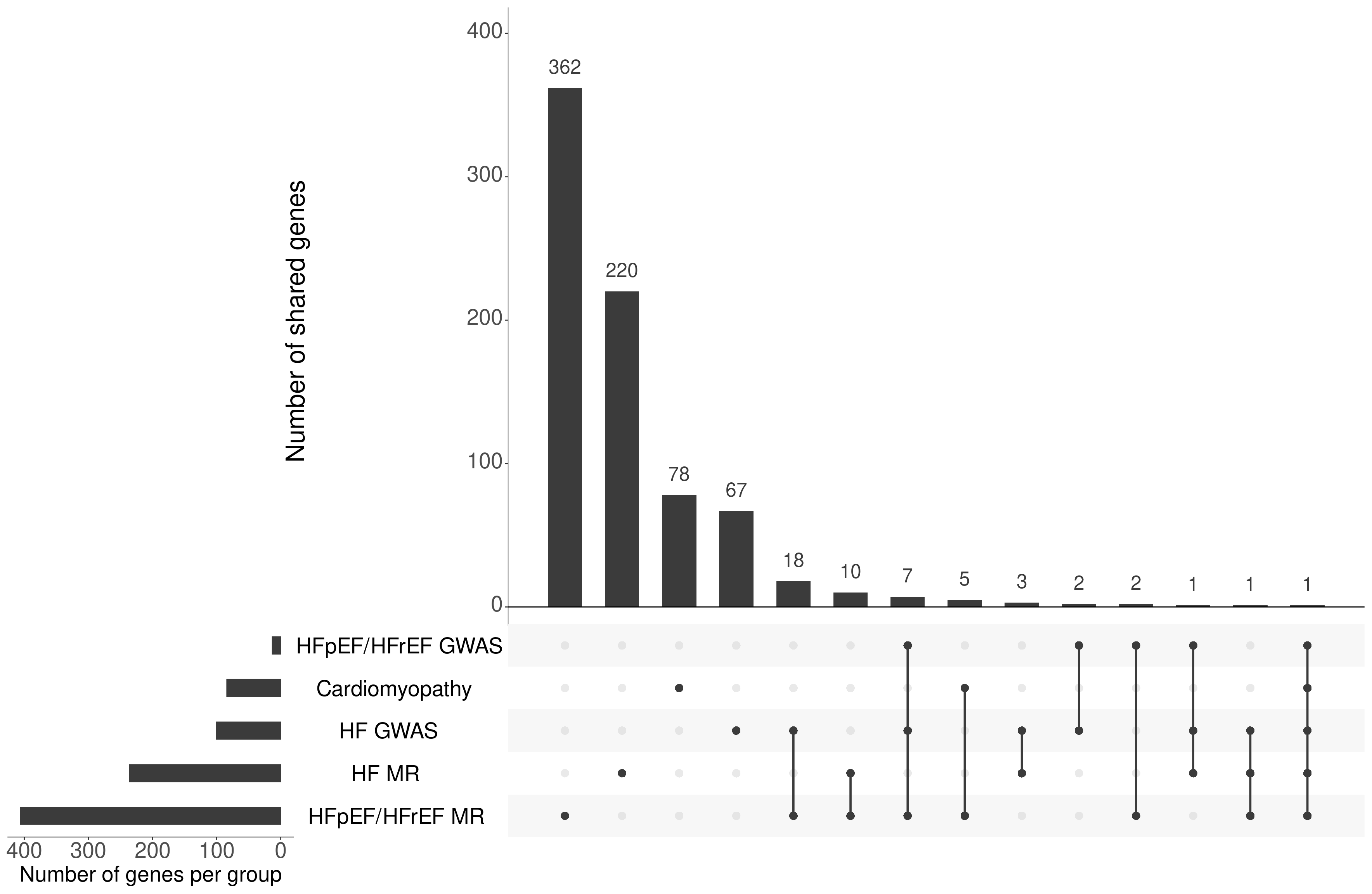

**Figure S2.** Venn diagram illustrating the overlap of genes among pQTLs **(Figure S2A)** and eQTLs **(Figure S2B)**. Figure 3A shows the overlap of genes among ARIC (1,342 genes), Fenland (1,383 genes), and deCODE (1,493 genes). Figure 3B shows the overlap of genes among eQTLGen (8,054 genes) and GTEx v8 (11,985 genes).

**(A)**

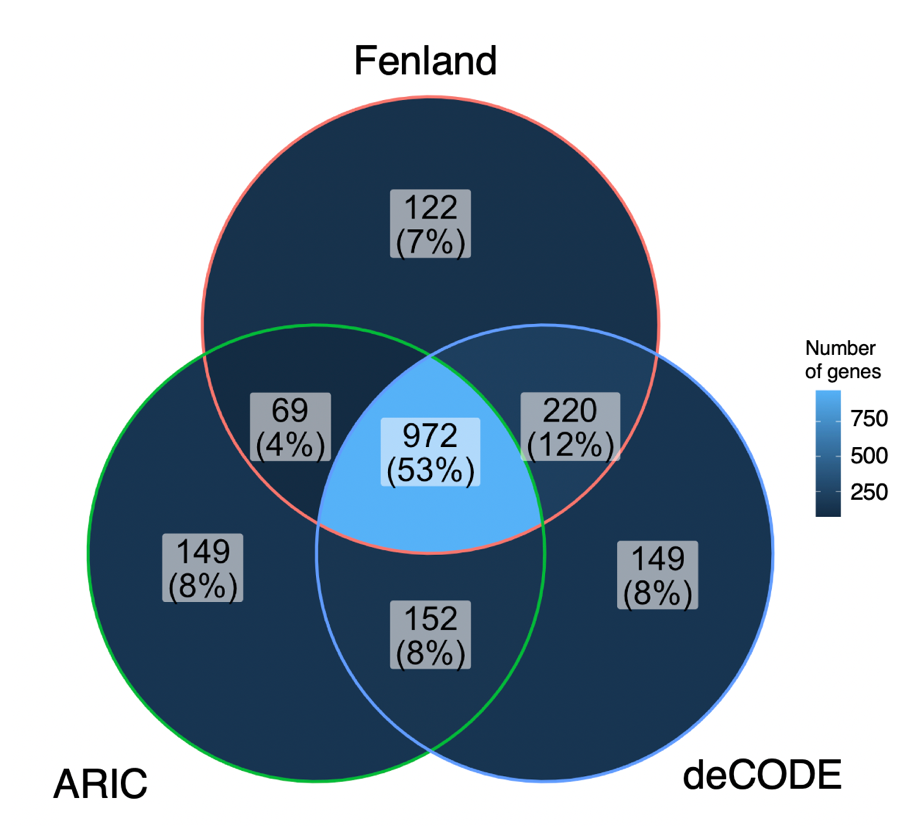

**(B)**

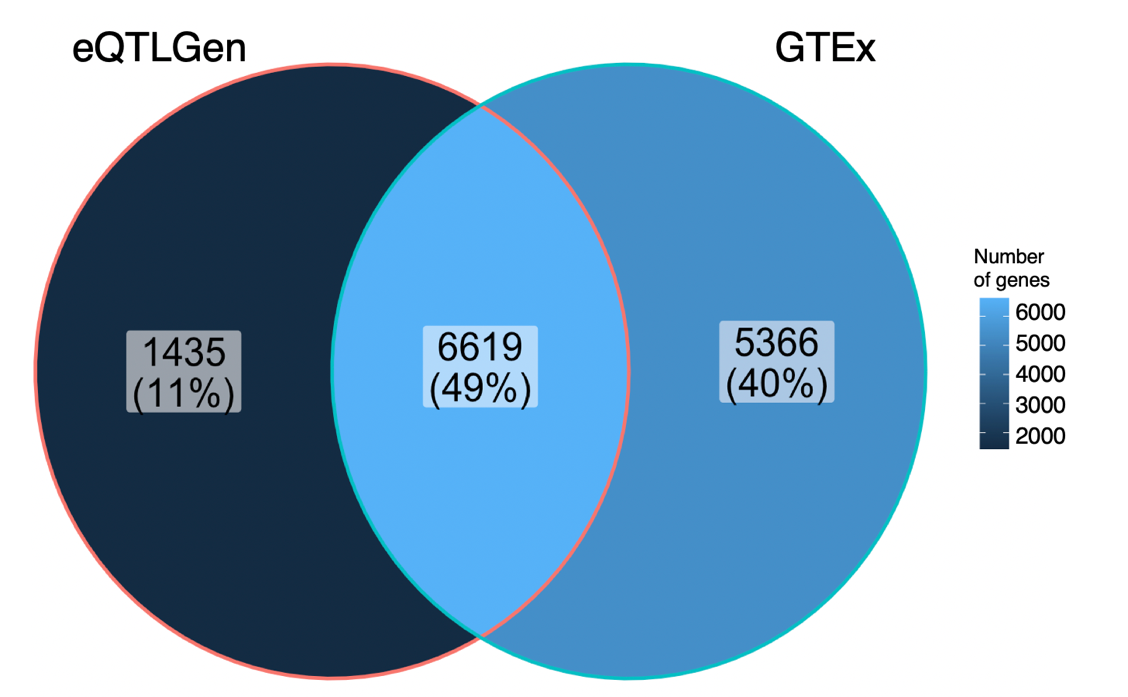

**Figure S3. Flowchart for druggability annotations of MR hits against HFrEF and HFpEF to identify potential repurposing and safety signals.**

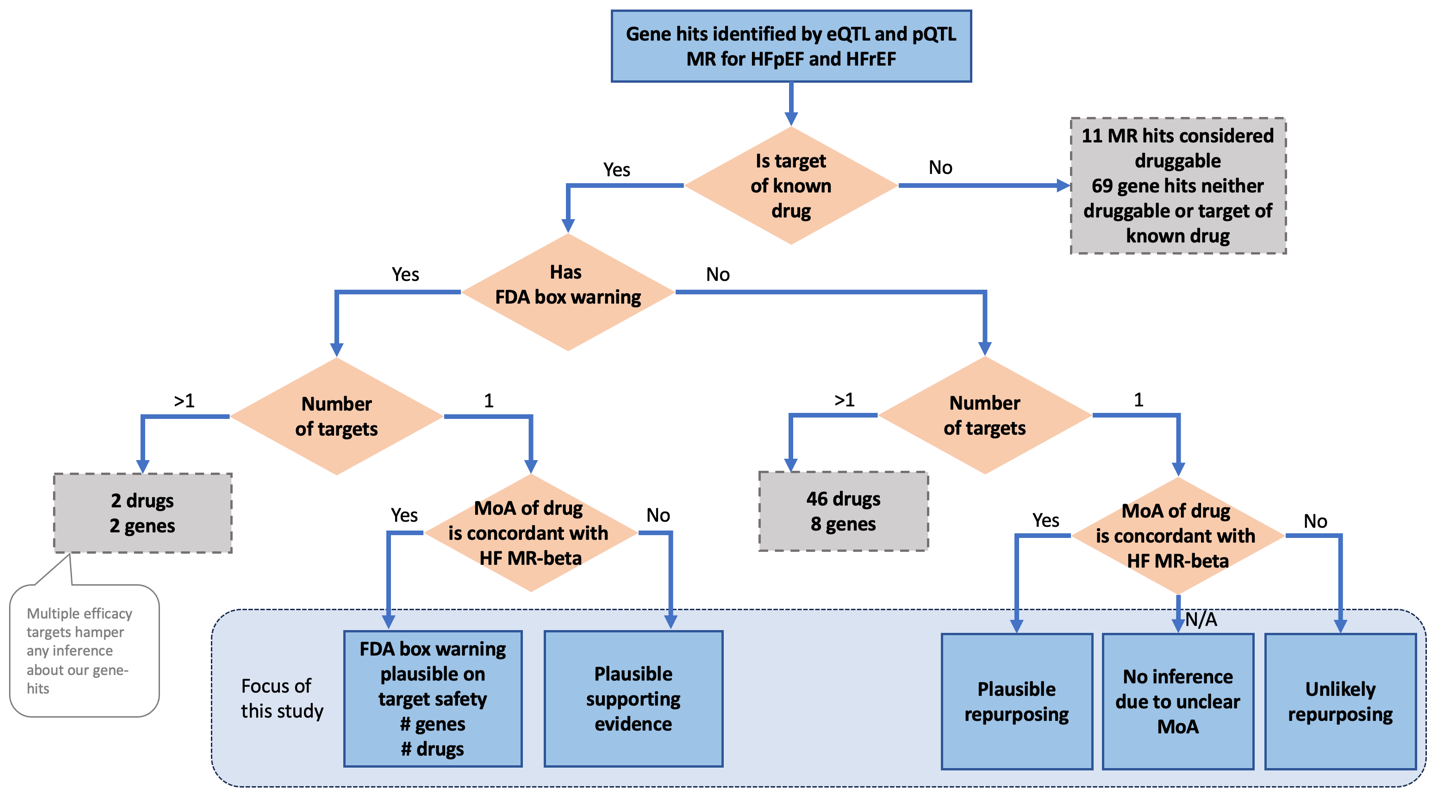

^i^Targets were efficacy targets obtained from ChEMBL version 32 (published 2/28/2023). This resource may be downloaded at <https://ftp.ebi.ac.uk/pub/databases/chembl/ChEMBLdb/releases/chembl_32/chembl_32_sqlite.tar.gz>.

Note: Data on >1 targets available on request; data on 1 target is presented in Supplementary Table on “Tractability”.
